# Biofilm Contributions of Bacterial Pathogens and Antimicrobial Resistance Genes to Wastewater Surveillance Signal at the Hospital Scale

**DOI:** 10.64898/2026.06.24.26356348

**Authors:** Amanda Darling, Sitara Sastry, Kate R. Bowie, Irvan Luhung, Andrew Franklin, Valerie J. Morley, Nicole Stephenson, Daniel Katz, Dawn Gratalo, Alexandra M. Simas, Taylor Burke, Madelena Ruedaflores, Scott C Roberts, Paul E. Turner, Richard A. Martinello, Jordan Peccia, Hannah G. Healy

## Abstract

Wastewater surveillance (WS) has been widely adopted as a cost-effective and population-representative infectious disease monitoring tool and is increasingly being applied to bacterial and antimicrobial resistance gene (ARG) targets. However, some of these targets may persist in pipe biofilms and detach into wastewater, complicating accurate WS interpretation. To investigate biofilm contributions to wastewater pathogen and ARG signals, paired sink-drain biofilm, branch-drain-plumbing biofilm (“sewer biofilm”), and wastewater were collected from five hospital sites over a four-month period and analyzed using 16S rRNA gene amplicon sequencing and probe-capture metagenomics. Overall, sewer biofilm bacterial communities were as diverse as wastewater. Across sites, a mean of 9% (0.9–23.3%) of wastewater bacterial communities could be attributed to sewer biofilm communities. Many clinically relevant pathogens were consistently detected both in sewer biofilm and wastewater, including environmentally persistent and/or biofilm-associated taxa (e.g., *Pseudomonas aeruginosa, Klebsiella pneumoniae)*. While many ARGs overlapped between wastewater and biofilms (e.g., *tetA, sul1, blaCTX-M, vanA*), others were significantly enriched in sewer biofilms (e.g., *qacL, van*-operon and *OXA* genes). Together, these findings confirm that wastewater pathogen and resistome profiles integrate inputs from both human shedding and pipe-resident communities and therefore need to be considered when selecting WS targets and interpreting signal.

**Synopsis:** Wastewater surveillance signal reflects bacteria from both human shedding and sewer-resident microbiomes. Paired biofilm and wastewater analyses demonstrated overlapping pathogens and resistomes, indicating pipe biofilms can shape signals used for population-level surveillance.

**TOC Abstract:** 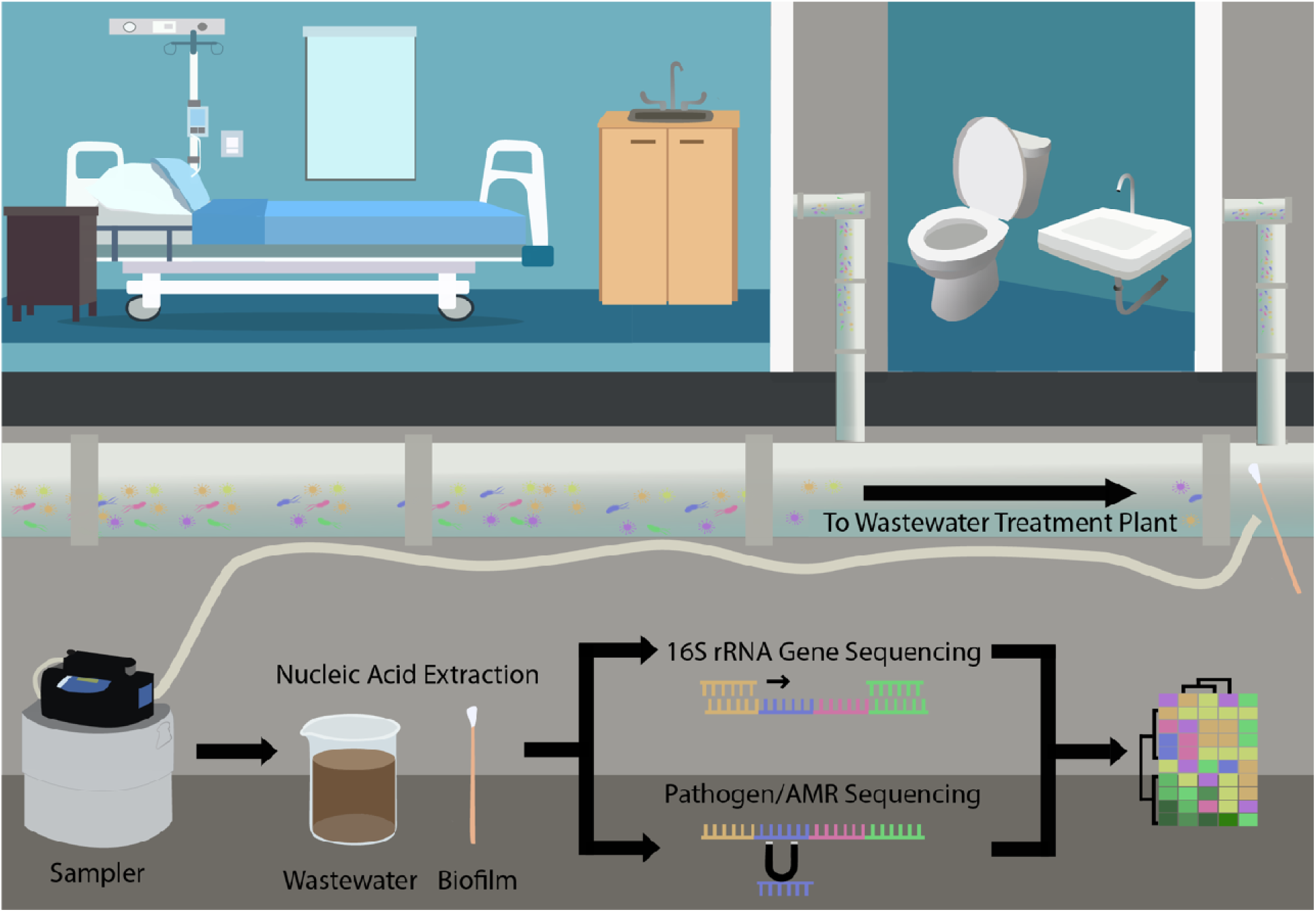

## INTRODUCTION

Wastewater surveillance (WS), the measurement of wastewater biomarkers to monitor community-level health within a defined sewershed,^1^ has been adopted widely to inform infectious disease dynamics. Applications of WS have included early warning of disease introduction, assessment of disease trends and seasonality, geographic hotspot identification, and detection of genomic variants.^2^ To date, WS has been validated at multiple scales (e.g., municipal, neighborhood, building) and for a growing list of communicable disease targets.^2^ Pathogens with the most immediate applicability for WS are (i) significant to public health, (ii) shed consistently in bodily fluids during infection such that biomarkers are detectable in wastewater, (iii) sufficiently stable in the environment, and (iv) lack the capacity for environmental growth outside a host.^3,4^ Viral pathogens that readily meet such criteria (e.g., SARS-CoV-2, RSV) have correspondingly comprised most WS applications thus far. However, extensions of WS are increasingly being proposed for monitoring bacterial pathogens (e.g., *Salmonella enterica*,^5^ fungal pathogens (e.g., *Candidozyma auris*^6^), and antimicrobial resistance gene (ARG) targets^7^ whose signals may be altered by in-sewer microbial activity. Such environmental contributions to wastewater pathogen and antimicrobial resistance (AMR) signals have been sparsely accounted for in WS frameworks, and the extent to which sewer biofilms can serve as reservoirs for WS-relevant pathogens remains unknown.

Prior amplicon sequencing-based characterizations of sewer microbiomes have estimated only 15-30% of wastewater bacterial communities are derived from human fecal sources,^8,9^ suggesting that the majority of bacterial communities in bulk wastewater originate elsewhere. Candidate non-human sources include environmental populations in sewer sediments and biofilms, tap water and stormwater, animal waste, and in-sewer growth in bulk wastewater.^10^ Among these, biofilms colonizing sewer pipe internal surfaces represent a particularly dynamic reservoir: their EPS matrix shields resident microorganisms from environmental stressors and facilitates ecological interactions such as nutrient scavenging and recycling.^11,12^ Biofilms are complex structures composed of microbial cells, primarily bacteria and archaea, embedded in a extracellular polymeric substance (EPS) matrix. In addition to serving as reservoirs for pathogens, biofilms are also likely AMR hotspots, as their EPS barrier confers reduced susceptibility to antimicrobials, they have a high cell-density, and can induce quorum sensing to synchronize horizontal gene transfer of ARGs.^13^ Through dispersal and sloughing, microorganisms can release from biofilms into bulk wastewater,^14^ potentially delivering pathogen signal downstream. Despite these posited contributions, the metagenomic resolution of biofilm-derived pathogen and ARG signatures in wastewater has not been explicitly studied *in situ* thus far.

WS for bacterial and fungal targets holds particular relevance in healthcare settings, given that many healthcare associated infections (HAIs) are caused by these agents (e.g., *Pseudomonas aeruginosa*, *Candidozyma auris*, *Klebsiella pneumoniae*), and hospital wastewaters have been identified as key reservoirs for such pathogens.^15^ The primary causative agents of HAIs include the ESKAPEE pathogens (i.e., *Enterococcus faecium, Staphylococcus aureus, Klebsiella pneumoniae, Acinetobacter baumannii, P. aeruginosa*, *Enterobacter* spp., and *Escherichia coli*).^16^ On average, 1 in 31 patients contract an HAI during their hospitalization.^17^ Further, HAIs are also at high risk of being drug-resistant, given an estimated 63.5% of drug-resistant infections are associated with healthcare.^18^ Potential drivers of drug-resistant HAIs include elevated antibiotic use in hospital settings, which selects for drug-resistant microbiota in human microbiomes.^19^ Further, excreted unmetabolized antibiotics in hospital plumbing can select for resistance in the environment.^20^ By characterizing HAI-associated pathogen and AMR dynamics within sewer systems, hospital-scale WS has the potential to transform outbreak response, guide antibiotic stewardship, and provide early warning of emerging resistance phenotypes.^21^

To inform WS target selection and interpretation, we aimed to characterize bacterial communities, pathogens, and ARGs present in plumbing biofilms and their potential contributions to wastewater signal. We collected samples of sink drain biofilms, tap water, sewer branch drain-pipe biofilms (“sewer biofilms”), and wastewater across five sites in a large, tertiary care hospital in the Northeastern United States over a 15-week period. Samples underwent 16S rRNA gene amplicon sequencing and hybridization-based probe-capture metagenomic sequencing to capture bacterial community, pathogen, and ARG profiles for each microbial compartment in hospital plumbing. Our primary objectives were to define the hospital plumbing microbiome and investigate its contribution to wastewater microbial signatures. To achieve this, we (1) characterized pipe-biofilm-associated microbial communities, (2) quantified how much of the wastewater bacterial community could be attributed to biofilm and tap water contributions, (3) identified pathogens and ARGs likely derived from sewer biofilms, and (4) assessed their spatiotemporal patterns.

## METHODS

### Study Location and Sample Collection

Sample collection was conducted across five sites in Yale New Haven Hospital (YNHH) in New Haven, CT, USA from August 27 to December 12, 2024. The five wastewater collection points represented diverse upstream populations and scales (**Table S1**). Site 1 captured approximately 55 toilets and sinks across Inpatient, General Surgery, Ambulatory Medicine, and Dialysis wards. Site 2 captured ∼31 patient bathrooms across Trauma Surgery and Pediatric Neurology wards and a Cardiac Intensive Care Unit (ICU). Site 3 collected from ∼49 bathrooms across Hematology Oncology, General Medicine, Infectious Disease, Inpatient, and General Surgery wards. Site 4 collected from 3 bathrooms across Cardiac and Surgical ICUs. Sites 1 and 3 were in the hospital’s East Pavillion, while Sites 2 and 4 were in the South Pavilion. Site 5 captured the entire North Pavilion, the Smilow Cancer Hospital, and represented ∼168 patient beds in addition to staff and visitor bathrooms, janitorial closets, hoppers, and handwashing sinks.

Wastewater samples were collected three times per week from each location using 24-hour composite CEC autosamplers (CA101, C.E.C. Analytics) connected to sewer-branch-drain pipes for each site. For Site 5, 1L of wastewater was collected on each sampling date with autosamplers set to run for 1 minute on, then 244 minutes off for 24 hours. Wastewater samples from all other locations were collected using continuous flow (run 1 minute on, then 1 minute off for 24 hours) into two 500 mL containers in series to enable collection of solids. Wastewater samples were transported to the lab on ice then aliquoted for downstream methods. Deviations from the prescribed sampling design (grab sampling events that represented a single time point of wastewater flow and composites collected over <24 hours) are described in **Table S2**. In total, 182 liquid wastewater samples were collected. Aliquots of wastewater samples for downstream 16S rRNA gene amplicon sequencing were stored frozen at −80°C prior to nucleic acid extraction, while wastewater aliqouts intended for probe-capture metagenomic sequencing were shipped on ice to Ceres Nanosciences for extraction and were never subject to freezing (Ceres Nanosciences, Inc., Manassas, VA).

Samples of hospital wastewater plumbing pipe biofilms (“sewer biofilm”) were collected from the same branch drains where wastewater sampling occurred. In addition, sink drain biofilms in fixtures upstream of each site’s wastewater collection point were sampled. Biofilm samples from sink drains and branch drains were collected at three experimental timepoints (immediately prior to, during, and shortly after the wastewater sampling period) (**Figure S1**). Biofilm samples were collected using BD Liquid Amies Elution Swab (“Eswab”) Collection/Transport System. For sink drains, swabs were inserted approximately 7.6 cm and rotated across the entire pipe circumference ten times (covering approx. 10 cm^2^). For sewer branch biofilms, swabs were inserted approximately 1.5 m into pipes via attachment to a wooden dowel, and the lower half circumference of the pipe was sampled by rotating the swab across it three times. Swabs were then reinserted into the Liquid Amies solution, sealed, and transported to the lab on ice to be processed within 24 hours. In total, 55 biofilm swabs (45 sink biofilm and 15 sewer biofilm) were collected and went forward for downstream processing.

Tap water samples were collected from fixtures upstream of Site 5 only (December 2023 to April 2024) as described in detail by Healy et al., 2025.^22^. Briefly, 2 L of first-flush tap water was collected in a Whirlpak® bag from each fixture (n=29), followed by sodium thiosulphate addition to quench chlorine. Following collection, samples were transported to the lab on ice then immediately filter-concentrated onto 0.2-μm PES filters (Millipore Sterivex). Filters were stored at 4 °C and then extracted for DNA within 24 hours.

### DNA Extraction

DNA extraction from biofilm swabs and drinking water filters were extracted according to a protocol previously described.^22^ Briefly, the filter cartridges were opened, cut with a sterile scalpel, and inserted into fresh tubes with lysis buffer, chloroform isoamyl alcohol, and beads followed by bead beating for 15 minutes. Alternatively, the swabs in Liquid Amies solution were bead-beaten for 15 minutes after adding lysis buffer and phenol-chloroform-isoamyl alcohol. The supernatant from both sample types was removed after centrifugation and added into a 24-well plate with proteinase K for automated extraction with a Kingfisher Apex using the MagMAX™ Microbiome Ultra Nucleic Acid Isolation Kit (Applied Biosystems). To recover nucleic acids from wastewater undergoing 16S sequencing, thawed 50 mL samples were centrifuged to concentrate solids followed by the addition of lysis buffer and phenol-chloroform-isoamyl alcohol to the pellet (mean 2.3 g). Samples were bead beaten for 15 minutes then centrifuged, and the supernatant was added into a 24-well plate with proteinase K for automated extraction with a Kingfisher Apex using the MagMAX™ Microbiome Ultra Nucleic Acid Isolation Kit (Applied Biosystems). After extraction of swabs and wastewater using the MagMAX kit, to remove RNA, RNase A was added to obtain a final concentration of 100 μg/mL and incubated at room temperature for 15 minutes prior to a column-based cleanup with the Genomic DNA Clean & Concentrator-10 kit (Zymo Research).

For wastewater aliquots subject to hybridization-based probe-capture metagenomic sequencing, samples (8 mL) were concentrated using Ceres Nanotrap Microbiome Particles (Series A and B) then extracted on a Kingfisher Apex using the MagMAX™ Microbiome Ultra Nucleic Acid Isolation Kit.

### 16S rRNA Amplicon Sequencing and Bioinformatics

Samples associated with three study timepoints (beginning, middle, and end) underwent 16S rRNA gene amplicon sequencing, including two wastewater samples per timepoint per site, three sink drain samples per timepoint per site, and one sewer branch sample per timepoint per site (**Table S3, Figure S1**). In addition, 29 tap water samples from Site 5 from December 2023-April 2024 were included. Samples were shipped on dry ice to the University of Minnesota Genomics Center (Minneapolis, MN, USA) for 16S-rRNA-gene amplicon library preparation and Illumina sequencing. Template DNA quantity in samples was assessed by creating 1:8 and 1:64 dilutions of each sample and subjecting them all to qPCR using primers V4F_Nextera and V4R_Nextera targeting the V4 locus in the 16S rRNA gene. Samples were diluted with nuclease-free water to normalize starting template volumes and to reduce the amount of PCR inhibitors present, and then amplicons were generated by subjecting sample DNA to either 25-cycle or 30-cycle PCRs (depending on how much template was found to be present in the samples during the initial qPCR) using the same primers. PCR products were diluted 1:100 in nuclease-free water and subjected to a second 10-cycle PCR to attach Illumina sequencing primer-compatible DNA regions as well as individual barcodes for each sample. Samples were all uniquely dual-indexed, as detailed in Gohl *et al*.^23^ Final PCR products were normalized using SequalPrep kits (Invitrogen), pooled into sequencing libraries, and cleaned with AMPure XP mag beads (Beckman Coulter). Sequencing libraries were loaded onto an Illumina MiSeq using a 2 × 300 v3 flow cell (Illumina, San Diego, CA, USA).

### Hybridization-Based Probe-Capture Metagenomic Sequencing

All wastewater samples underwent hybridization-based probe-capture metagenomic sequencing (“probe-capture metagenomics”) Illumina®/IDbyDNA Respiratory Pathogen ID/AMR Panel (RPIP) that enriches for 180+ bacteria, 50+ fungi, and 40+ viruses and antimicrobial resistance alleles (1200+) (**Table S4**). A set of sewer biofilm samples from each site at three timepoints (beginning, middle, end) as well as three sink drain biofilm samples per site at the end timepoint were also sequenced using this approach (**Figure S1**, **Table S3**). Libraries were sequenced using the Illumina NovaSeq 6000. All sequencing runs included no template controls (NTCs) and positive viral controls. The raw sequence reads used in this study can be found in the Sequence Read Archive (SRA) under BioProject ID: PRJNA1469170.

### Data Analysis

#### Bioinformatics for 16S rRNA Amplicon Sequencing Analysis and Source Tracking

The 16S rRNA gene amplicon sequences were curated using the DADA2 pipeline (v1.36) and phyloseq (v1.52) in R. Taxonomy classification of amplicon sequence variants (ASVs) was conducted using the Silva database (v138.2; https://zenodo.org/records/14169026). Decontamination and differential gene abundance analysis were performed using DESeq2 (v1.48.1). Reads below 1,000 were filtered out in decontamination steps. Rarefaction curves for all samples plateaued, suggesting sufficient sequencing depth (**Figure S2**); therefore, rarefaction was not performed. We employed fast-expectation-maximization microbial source tracking (FEAST) to estimate the source attributions from biofilm and tap water to downstream bulk wastewater microbiomes using 16S rRNA-gene amplicon sequencing data. FEAST (FEAST v0.1.0)^24^ analyses were performed for 3 timepoints for each of the 5 site locations. All tap water samples were used as sources for each timepoint and site. Tap water, sink biofilm, and sewer biofilm samples were designated as sources while wastewater samples were designated as sinks. We also examined the number of ASVs shared between sample types by class (ggupset v 0.4.1).

#### Bioinformatics for Probe-Capture Metagenomic Sequencing

Adaptors were trimmed using fastp v0.23.4, and human reads were removed using custom database built on https://www.ncbi.nlm.nih.gov/datasets/genome/GCF_000001405.39/ (Kraken2 v2.1.2). Filtered reads were then annotated using Kraken 2 v2.1.3 for general microbial taxonomy and community structure against a custom Kraken2 database built from RefSeq bacterial, viral, plasmid, and fungal libraries (built January 2025, Kraken2 v2.1.3) and CARD v4.0.1 for ARGs. Analysis of the probe-capture metagenomics dataset was limited to a predefined list of pathogen targets included in the RPIP panel (**Table S4**). For Kraken2, we set a confidence threshold of 0.5 and a minimum number of 3 k-mer hit groups to limit spurious annotations. Relative abundances were calculated from Kraken2 taxonomic classification reports. For each sample, species-level read counts were normalized by the total number of quality-filtered, host-depleted reads passing into Kraken2, as determined from FastQC reports run on trimmed and de-hosted reads. Only pathogens detected above a relative abundance of 0.0001% were retained for analyses to limit spurious detections. We also integrated a qualitative check on Kraken2 annotated reads described in detail in the **Supplemental Text**. Briefly, we used BLASTn to confirm alignment for each sample-taxid pair identified. For CARD annotation, the blastx DIAMOND (version: 0.9.24.125) function was used with filtering for a minimum identity of 80%, minimum length of 25, and a minimum e-value of 1 × 10^−10^. To holistically capture all ARGs relevant to selection pressures in sewer environments (e.g., genes conferring resistance to quaternary ammonium compounds), we did not filter the probe-capture metagenomic dataset to ARG families exclusively enriched in the panel. ARG abundances were normalized as reads per kilobase per million mapped reads (RPKM), while taxonomic relative abundances were reported without normalization.

As a sensitivity analysis to ensure pathogen detections were not overly conservative or permissive, we also annotated reads using MetaPhlAn v4.2.2 for pathogen species markers, Sylph v0.9.0 against the GDTB-R220 database for low abundance bacterial taxa, and Centrifuger v1.0.12 against the pre-built RefSeq-based database of human, bacterial, archaeal, viral, and SARS-CoV-2 genomes (cfr_hpv+gbsarscov2.1–3.cfr),^25^ downloaded from Zenodo for prokaryotic and viral genomes. Databases and bioinformatics pipelines for metagenomic analyses are available at https://github.com/avdarlingphd/darling-2026-biofilm-wbe.

#### Statistical Analyses

Plots and analyses were generated in RStudio, R version 4.5.1, using the tidyverse (v2.0.0)^26^, ggplot2 (v4.0.2)^27^, phyloseq (v1.54.2)^28^, vegan (v2.7.3)^29^ and dplyr (v1.2.0)^30^ packages. Nonmetric multidimensional scaling (NMDS) plots were generated using the metaMDS function in vegan. Pairwise univariate differences between two groups were assessed using the Wilcoxon rank-sum test, while Dunn’s test was employed post hoc for comparisons among three or more groups. Where applicable, p-values were adjusted for multiple comparison using the Bonferroni procedure. Alpha diversity was quantified using the Shannon index. Differences in community composition and dispersion were evaluated using PERMANOVA on Bray-Curtis-transformed distance matrices for 16S rRNA amplicon sequencing and probe-capture metagenomic datasets. MaAslin3 was used to evaluate differentially abundant pathogens, ARGs, and ARG drug classes between sewer biofilm and wastewater. The fixed effect was the sample matrix distinction (sewer biofilm vs wastewater) whereas respective hospital sampling sites were set as random effects. For all analyses using RPIP data where sample types were compared, we retained at most one hospital wastewater sample per biofilm-collection timepoint (timepoint 1: August 27–September 12, timepoint 2: October 28–31, and timepoint 3: December 4-12), selected as the sample closest in collection date to the corresponding biofilm sample collection (**Figure S1**).

## RESULTS AND DISCUSSION

Over the study period, samples collected across five hospital sites (**Table S3, Figure S1**) included 181 wastewater samples, 15 sewer plumbing branch drain biofilm (“sewer biofilm”) samples, and 45 sink-drain pipe biofilm (“sink biofilm”) samples, as well as 29 tap water samples from Site 5. Following quality control and filtering, 209 of 210 samples that underwent probe-capture metagenomic sequencing with the Respiratory Pathogen ID/AMR (RPIP) panel (181 wastewater, 15 sewer biofilm, 13 sink drain biofilm) and 118 of 119 samples that underwent 16S rRNA gene amplicon sequencing (30 wastewater, 14 sewer biofilm, 45 sink drain biofilm, 29 tap water) met minimum read thresholds for downstream analyses (**Table S3, Table S5**). The total number of reads that passed quality control, dehosting, and trimming steps alongside the total number of reads annotated by Kraken2 are provided in **Table S6**. Deviations from sampling protocols and reasons for sample-specific sequencing failures are provided in **Table S2**. The average number of reads per sample was 30.2 million for RPIP metagenomes and 70,400 for 16S amplicon libraries. For the probe-capture metagenomic dataset, 26.4% of total reads classified using Kraken2 were assigned to pathogens included in the RPIP panel; the remaining taxa were excluded from the analysis. Among detected sample–pathogen combinations, the mean fraction of the reference genome covered at ≥1x was 0.6% (methodological details in the **Supplemental Text**), likely reflecting probe-based enrichment of the targeted regions of pathogen genomes.

### Core Microbiome of Sewer Biofilm

At the family level, bacterial communities were largely comprised of *Comamonadaceae, Propionibacteriaceae, Rhodocylcaceae,* and *Microbacteriaceae* (**Figure 1a**). Several genera met our criteria for inclusion in the sewer biofilm “core” community (≥75% sample prevalence at ≥0.25% relative abundance): *Acidovorax, Leucobacter, Microbacterium, Propioniciclava, Tessaracoccus, Flavobacterium, Chryseobacterium, Cloacibacterium, TM7a, Bosea, Shinella, Comamonas, Ottowia, Aeromonas,* and *Stenotrophomonas* (**Figure 1c**). Though literature characterizing sewer biofilm core microbiota is limited, several of these genera were reported at abundant levels in hospital sewer biofilms grown for seven days on polystyrene coupons (*Acidovorax*, *Aeromonas, Stenotrophomonas, Flavobacterium, Comamonas*)^12^. However, other genera previously described as dominant in plumbing biofilm (*Pseudomonas, Acinetobacter, Arcobacter*)^12,31^ did not meet our ‘core’ definition due to presence at lower abundances **(Figure 1c).** The genera with the highest mean relative abundance in sewer biofilms across sites were *Flavobacterium* (1.32-15.0%, mean:7.9%), *Propioniciclava* (0.77-17.7%, mean: 5.67%), *Acidovorax* (3.00-11.8%, mean: 5.55%), and *Cloacibacterium* (0.82-9.76%; mean: 4.74%) (**Figure S3**). Sewer biofilm composition appeared to be characterized by a taxonomically diverse and flexible assemblage, consistent with biofilms responding dynamically to localized plumbing conditions.^32^ Combined, these findings highlight individual genera and families in consistently high abundance that may contribute to biofilm-derived wastewater signals.

**Figure 1.**
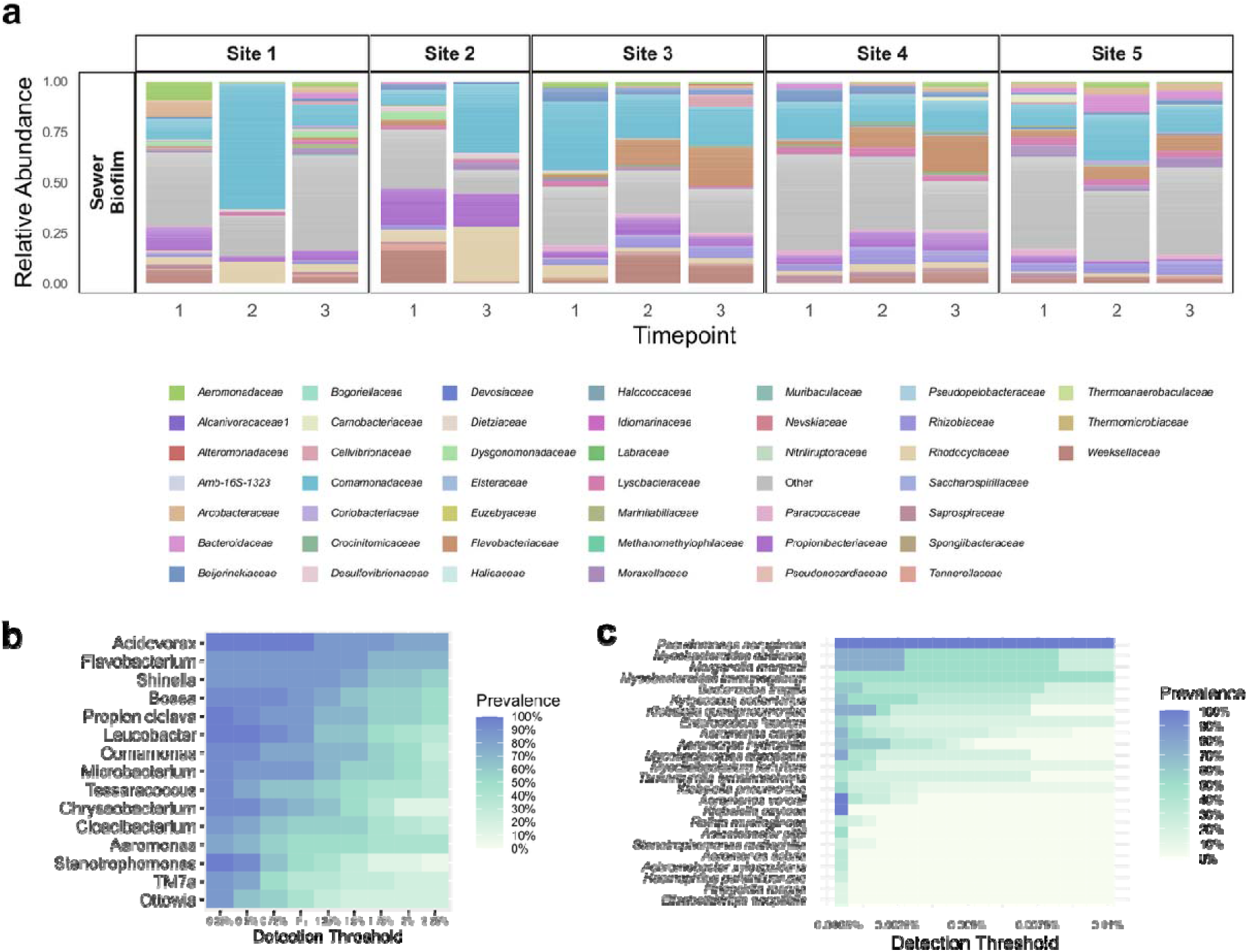
**a) Relative abundance of 16S-derived families in sewer biofilm samples across sites and timepoint.** Stacked bars show the top 44 genera by mean relative abundance; each bar is a unique timepoint–date sampling event. **b) Core microbiome (16S).** Heatmap shows the proportion of genera with ≥75% sample prevalence at ≥0.25% relative abundance. **c) Core pathogens across detection thresholds.** Heatmap shows prevalence (%) of RPIP-listed pathogens in sewer biofilm samples across relative abundance thresholds (0.0005%–0.01%), where prevalence is the percentage of samples exceeding each threshold. Only pathogens with ≥10% sample prevalence at ≥0.0005% relative abundance were included.

Sewer biofilm pathogen profiles were composed of species with documented biofilm-association, though to varying degrees. Among enriched pathogens (**Table S4**), those detected at higher relative abundances in sewer biofilm (≥10% sample prevalence at ≥0.0005% relative abundance) included *P. aeruginosa*, *Klebsiella pneumoniae*, *Enterococcus faecium, Aeromonas hydrophila*, and *Aeromonas caviae* (**Figure 1d**, **Figure 3a**). Largely, pathogens above this abundance threshold in sewer biofilms were those with previously documented association with biofilms, with some specifically documented in plumbing biofilms (*P. aeruginosa, S. maltophilia, K. pneumoniae, E. faecium*) ^33–35^. Some pathogens with literature-supported sustained presence in sewer plumbing biofilm, including *E. coli* and *Proteus mirabilis* ^34^, however, were only present below this threshold (**Figure 3a**, **Figure 1d).** *P. aeruginosa* was the most abundant pathogen in the sewer biofilm community (**Figure 1d)** despite the absence of *Psuedomonas* spp. from the 16S-derived core microbiome (**Figure 1c**), likely reflecting successful enrichment of *P. aeruginosa* by the probe-capture panel. Overall, the presence of these pathogenic taxa both underscores the potential role of sewer biofilms as persistent reservoirs for pathogen signal contributions and delineates which species present planktonically in wastewater might likely be sourced from such reservoirs.

### Community Sharedness and Source Contributions of Biofilm Bacterial Communities to Wastewater

Microbiome diversity increased across environments (tap water < sink biofilm < wastewater and sewer biofilm), consistent with cumulative microbial inputs along building plumbing gradients^36^ (**Figure 2a, Table S7, Table S8**). Although NMDS ordination showed partial overlap between sewer biofilm and wastewater communities, pairwise PERMANOVA (p<0.05) comparison indicated that community composition differed significantly, suggesting that while taxa were shared, their relative abundances or overall structure were distinct (**Figure 2b, Table S7, Table S8)**. Variation in community composition was primarily explained by environment (22.7%), followed by site (13.5%) and the interaction between environment and site (11.4%). These results indicate that bacterial community structure is spatially heterogeneous, shaped both by environmental niche and local conditions in the pipe network. Broadly, these findings indicate that sewer biofilms represent a microbially diverse and compositionally distinct environment, with potential to shape wastewater-associated microbial signals.

**Figure 2.**
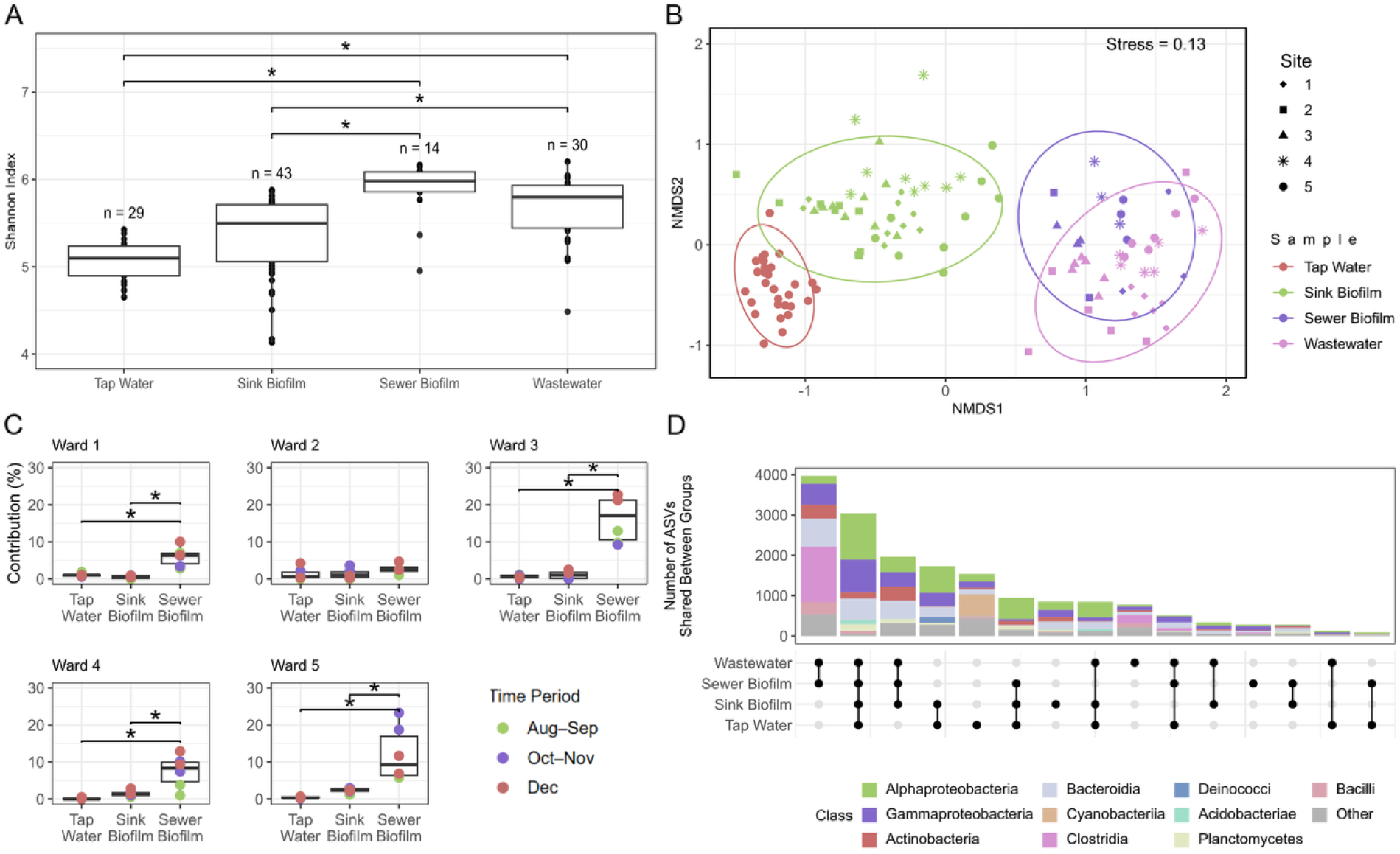
16S rRNA amplicon sequencing results. a) Shannon diversity index by sample type. b) Non-metri Multidimensional Scaling (NMDS) plot by sample type and site with stress 0.13. c) Fast expectation-maximization microbial source tracking (FEAST) percent contribution to wastewater by site. d) Shared Amplicon Sequence Variants (ASVs) between sample types by Class. Brackets and asterisks on box plots indicate significant differences between sample types (p<0.05). Box edges correspond to the first quartile, median, and third quartile. Whiskers range from the edge to the largest and smallest values.

Although sewer biofilm and wastewater communities were compositionally distinct (PERMANOVA, p<0.05), 44% of detected bacterial genera (n = 288) were shared between the two environments (**Figure 2d, Figure S4**). Of the tested environments, sewer biofilms also contributed a larger share of bacterial community signal to wastewater (0.9–23.3% of wastewater bacterial community composition; mean: 9%; FEAST) compared with sink biofilms (<0.001-0.04 %; mean: 1.3%) or tap water (<0.001-0.04 %; mean: 0.7%) (**Figure 1c**). At the amplicon sequencing variant (ASV) level, most ASVs were detected in exclusively both sewer biofilm and wastewater at least once (**Figure 2d**). These observed overlaps in bacterial community composition, as well as source attribution of wastewater bacterial signatures to sewer biofilm, suggest ongoing exchange of microbial taxa between both environment.^14,37^

### Overlapping Pathogen Detection in Wastewater and Biofilms

Consistent overlap of pathogenic species was observed between sewer biofilm and wastewater metagenomes, with probe-capture metagenomes between the two environments significantly similar in composition (Pairwise PERMANOVA p<0.05) (**Table S9, Figure S5, Figure S6, Figure 3a, Figure S7, Figure S8**). Wastewater and sewer biofilm, however, had significantly higher diversity of pathogens compared to sink biofilm (**Figure S6**). Though fungal pathogens (*Fusarium verticillioides, Fusarium oxysporum*) were detected in some samples (n = 4), detected pathogens were largely bacterial (**Figure 3a**). Pathogens detected consistently in all evaluated environments (sink biofilm, sewer biofilm, and wastewater) included *P. aeruginosa, Klebsiella pneumoniae,* and *Stenotrophomonas maltophilia* (**Figure 3a**). The majority of detected pathogens (n=41; 52%) were shared between sewer biofilm and wastewater (**Figure S8, Figure S9, Figure S10)**. Of these, many were biofilm-associated pathogens linked to both human and aquatic reservoirs, including *Acinetobacter* spp., *Aeromonas* spp., *P. aeruginosa*, *S. maltophilia,* and *Klebsiella* spp.^38^ Additionally, several pathogens belonging to the family *Enterobacteriaceae* — whose pathogens have been isolated previously from hospital plumbing at clonal identities unrelated to local infection ^39^ — were shared between biofilm and wastewater including *E. coli, Klebsiella* spp. (*K. quasipneumoniae, K. oxytoca, K. variicola, K. aerogenes), Raoultella ornithinolytica, Morganella morganii, Proteus mirabilis,* and *Serratia marcescens*. Several additional species whose primary reservoir is the human gastrointestinal or urinary tract (e.g., *Bacteroides fragilis*, *Enterococcus faecium*, *Morganella morganii*, and, under the classical view, *E. coli*) were also detected in both environments, likely reflecting transient accumulation of human-derived signals in pipe biofilm and/or loose deposits.^37^ Pathogens significantly enriched in sewer biofilm over wastewater also included those with both human and aquatic reservoirs (*A. baumannii, A. caviae*, *A. veronii*) (MaAslin3, p<0.05) (**Figure S11**), suggesting biofilm rather than active infection as a likely wastewater signal source. These species, along with *P. aeruginosa, K. pneumoniae,* and *A. hydrophila —*detected in at least three sewer biofilm samples and at least 170 wastewater samples **(Figure S12)—** are thus expected to consistently overlap in both environments. Of those shared between sink biofilm and wastewater, the large majority were oral commensals or opportunists consistent with transient human-derived inputs to environmental biofilms (e.g., *Actinomyces israelii*, *Gemella haemolysans*, *G. morbillorum*, *Neisseria mucosa*, *Streptococcus intermedius*), along with the gut anaerobe *Clostridium perfringens*, whose detection likely reflects spore-mediated persistence rather than active biofilm growth^40^ (**Figure 3a**). Together, these findings highlight the overlap in pathogenic species between biofilms and wastewater and the potential for exchange of microorganisms between the two environments.

**Figure 3.**
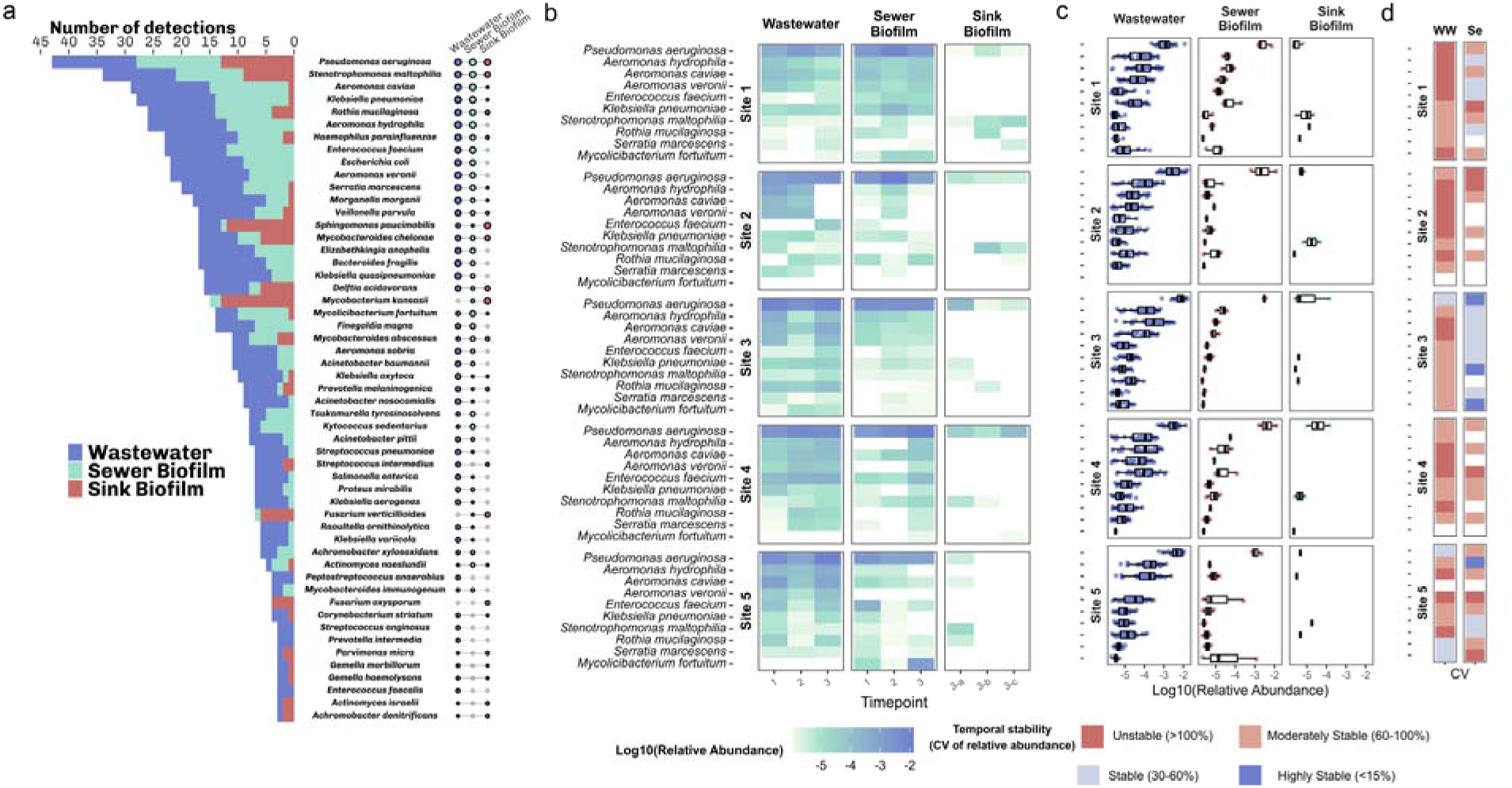
**a) Detection frequency of RPIP pathogen species across sample types.** Bars show the number of samples in which each species was detected across hospital sites. The accompanying dot plot is scaled to detections within each type. Only RPIP target species detected at least twice are included. **b) Relative abundance heatmaps of the top RPIP pathogen species by site across sample types.** Top 10 species were selected by median relative abundance across sewer biofilm sampling dates. Panels are faceted by site (rows) and sample type (columns: sink biofilm, sewer biofilm, wastewater), with timepoints on the x-axis; only wastewater dates shared with biofilm collections are shown. Color intensity represents log□□-transformed relative abundance from Kraken2 classifications. **c) Relative abundance distributions.** Box plots show log□□-transformed relative abundance for the same pathogens across sample types and sites (points = samples, boxes = IQR, line = median). Pathogens are ordered as in panel b; empty space indicates non-detection for a given site–sample type combination. **d) Coefficient of variation (CV) heatmap.** Temporal stability of the top 10 pathogens, with CV of relative abundance calculated across sampling dates within each site and sample type and binned into stability classes (highly stable to highly unstable). Only pathogens detected in more than one sample type within a site are included; facets are sites (stacked vertically), with sample types on the x-axis. Pathogens are ordered globally by decreasing median relative abundance in sewer biofilm. White in panels a–c indicates non-detects; CVs are shown only for wastewater (WW) and sewer biofilm (Se).

### Biofilm-associated pathogen signals in wastewater were consistent over time

Several pathogens shared between biofilm and wastewater were detected consistently across sampling events and environments, indicating sustained presence rather than sporadic introduction. Notably, *Pseudomonas aeruginosa*, whose genera are known permanent residents of sewer system biofilms,^37^ exhibited the highest median relative abundance across sites, with moderate temporal variability in wastewater (coefficient of variation (CV) < 60% for 2/5 sites) (**Figure 3c**, **Figure 3d**). *P. aeruginosa* was also the dominant pathogen compositionally in both wastewater (0.05-1.26%, mean: 0.46%) and sewer biofilm (0.06-1.26%, mean: 0.38%) for all sites (**Figure S13**). Comparatively abundant pathogens in sewer biofilms included *A. caviae*, *Enterococcus faecium*, *A. hydrophila*, *S. maltophilia*, and *K. pneumoniae* (**Figure 3c**). The sustained presence of these pathogens supports in-sewer persistence and suggests that biofilm-derived signals may be systematically accounted for or excluded using correction factors in WS frameworks. Establishing baseline pathogen loads would be critical for developing such correction approaches.

### A subset of bacterial pathogens was detected exclusively in wastewater and not in paired biofilm samples

Of the detected pathogens, 27% (n=22) were observed only in wastewater (**Figure S8**). This group included several bacterial pathogens previously proposed as suitable WS targets: *Bordetella* spp. (*B. pertussis*, *B. parapertussis, B. bronchiseptica*),^41^ *Streptococcus* spp. (*S. anginosus, S. mitis*)^42^ (**Figure 3a, Figure S7**). However, *Enterococcus faecalis*, a well-characterized biofilm former in clinical contexts^43^ also met this distinction. Together, these results suggest that wastewater-only detection can help confirm or identify candidate WS targets. However, absence from sampled biofilms should be interpreted cautiously for some targets (*E. faecalis*), given inherent heterogeneity in biofilms, deposition & resuspension dynamics, and a relatively small sample size.

### Bioinformatic Workflow Shaped Observed Pathogen Profiles

We identified markedly different pathogen profiles when applying four commonly used taxonomic classifiers: Sylph (v0.9.0), Centrifuger (v1.0.12), MetaPhlAn (v4.2.2), and Kraken2 (v2.17.1) with varying confidence thresholds. Given the diversity of available taxonomic profilers,^44–46^ and ongoing challenges identifying low-abundance pathogen reads obscured by host and mammalian background,^47^ bioinformatic method selection is critical for WS. This study enabled qualitative validation of classifier output against both clinical context, provided by consulted hospital clinicians, as well as culture-based context specific to the sink biofilms sampled detailed in previous work.^22^ We also confirmed whether classified reads were false positives using BLASTn (**Supplemental Text**). We found that Kraken2, when applied with our chosen quality control parameters (confidence threshold of 0.5 and a relative abundance threshold of 0.0001%), yielded the appropriate balance of precision and recall for our study by detecting pathogens known to persist in sink drain biofilms without introducing false positives— rare pathogens with no clinical detection that were not confirmed with BLASTn (e.g., *Yersinia pestis* and *Bacillus anthracis)*. In contrast, MetaPhlAn and Sylph produced overly conservative pathogen profiles. Marker-gene-based approaches like MetaPhlAn, however, likely were not suitable for probe-capture metagenomics, since the designed probes may not capture the MetaPhlAn-delineated marker genes. More permissive approaches (Centrifuge/r and Kraken2 at lower confidence thresholds of 0 and 0.2) produced expanded pathogen profiles, including likely false positives (**Figure 3a, Figure S14, Figure S15, Figure S16, Figure S17, Figure S18**). While such false positives can be readily identified in a controlled hospital setting, misinterpretation in less-equipped settings where WS is applied could erroneously signal outbreaks and misinform public health responses. As sequencing methodologies and metagenomic approaches for WS continue to evolve, interpretation of pathogen profiles must similarly evolve, build toward methodological standardization, and be guided by consultation with subject-matter experts in infectious disease and environmental microbiology.

### Biofilm Contributions of Antimicrobial Resistance Genes to Wastewater

In addition to bacterial pathogens, biofilms can serve as reservoirs for antimicrobial resistance genes carried by pathogenic and nonpathogenic organisms or maintained within mobile genetic elements or extracellular DNA in the biofilm matrix.^48,49^ Though resistome composition and alpha diversity of ARGs were significantly distinct between environments (PERMANOVA, p<0.05, **Table S10, Figure S19, Figure S20),** ARG overlap between sewer biofilm and wastewater was substantial. A total of 1,568 (66% of total detected ARGs) ARGs were detected in both sewer biofilm and wastewater **(Figure S21**). A total of 1,046 ARGs were detected and shared exclusively in sewer biofilm and wastewater, exceeding the number detected exclusively in hospital wastewater (n = 550), further indicating substantial resistome overlap (**Figure 4b)**. Several ARGs that have been proposed as quantitative indicators of AMR proliferation in water and wastewater (e.g., *tet*A, *sul*1, *bla*CTX-M, *van*A)^50,51^ were detected in all environments (**Figure 4a**). Correspondingly, most resistance drug classes were detected in all environments (**Figure S22, Figure S23**). This widespread occurrence of ARGs in plumbing biofilms suggests that ARG detection in wastewater likely reflects a combination of human shedding, in-sewer selection, and biofilm-derived inputs. Consistent with this, prior work has documented how ARGs tend to be enriched in biofilms.^52,53^ We also detected rare genes of clinical concern in sewer biofilms (*mcr* 5.1, *mec*A),^51^ signaling potential presence of multidrug- and pan-drug-resistant pathogens that persist in-sewer. Combined, these findings highlight how ARG flux in-sewer complicates straightforward interpretation of wastewater data for tracking population-level AMR circulation, but WS still confers important One Health benefits, such as identifying emerging resistant strains.^54^ As an alternative framework, WS could be leveraged to establish baseline resistome signatures, enabling detection of deviations from baseline ARG profiles as indicators of emerging resistance or shifts in selective pressures.

**Figure 4.**
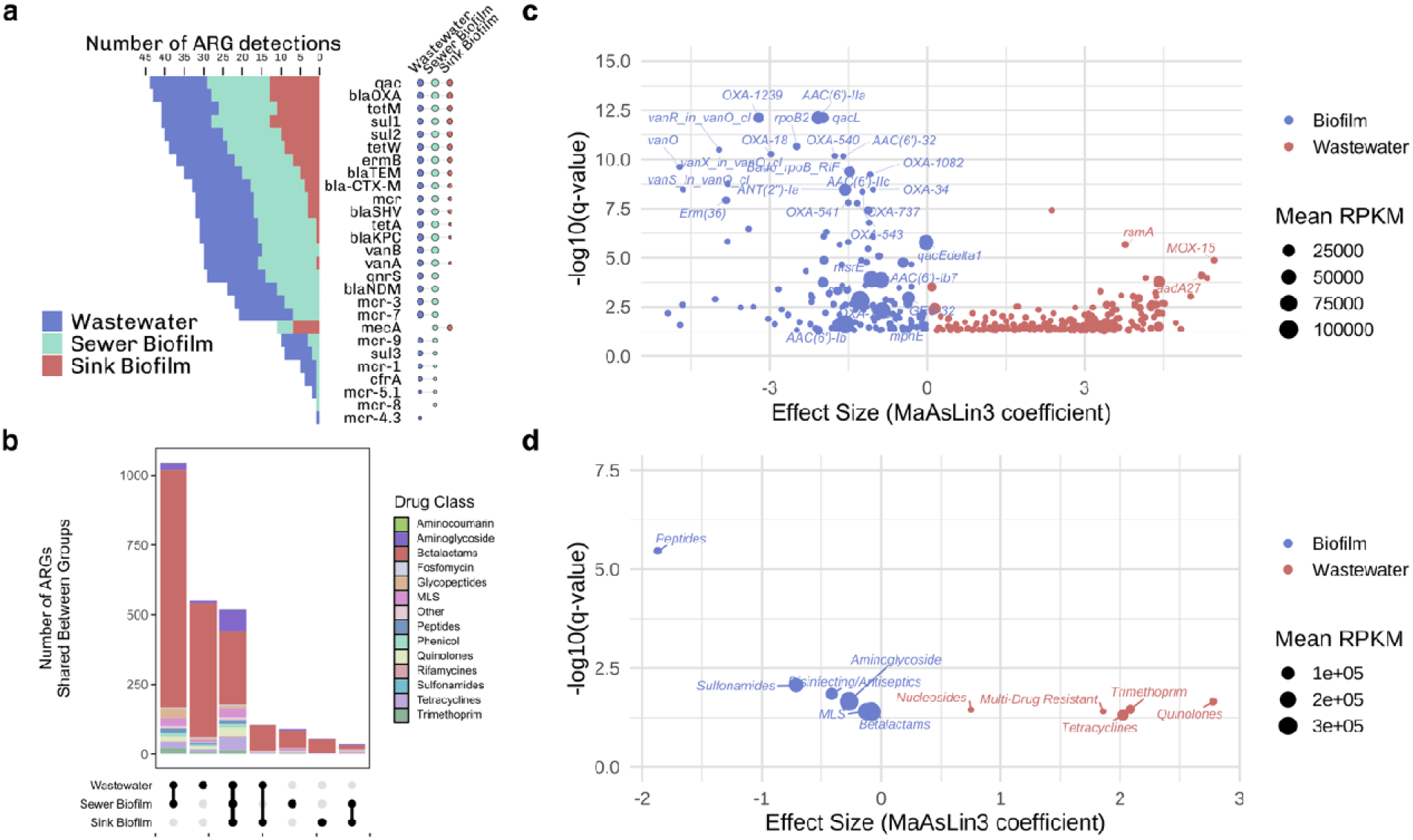
**a) Detection frequency of indicator ARGs across sample types.** Bars show the number of samples in which each ARG was detected at least once across sites. The accompanying dot plot shows detection distribution, with dot size scaled to detections within each type. **b) UpSet plots of ARG overlap across sink biofilm, sewer biofilm, and hospital wastewater, faceted by site.** ARG detections were derived from CARD profiles and binarize (present if detected in any sample from a given environment). Bars indicate the number of ARGs in each sample-typ combination, colored by Drug Class (top-ranked classes shown; remainder grouped as “Other”). **c–d) Volcano plot comparing abundance in sewer biofilm vs. bulk wastewater for c) ARGs and d) ARG drug classes.** Positiv effect sizes indicate higher abundance in bulk wastewater; negative indicate higher abundance in sewer biofilm. Point size reflects mean relative abundance across samples, and the genes/drug classes with the lowest p-values are labeled.

Some ARGs were differentially enriched in either sewer biofilms or wastewater (**Figure 4c**). ARGs enriched in biofilm aligned with expected selection pressures present in pipe environments (e.g., disinfectants and other antimicrobials). These included genes that are associated with biofilm-forming, environmentally persistent, opportunistic pathogens (e.g., van operon genes in *Enterococcus* spp., OXAs in *Acinetobacter baumannii* and *Enterobacteriaceae*^55^) and that confer resistance to biocides applied to surfaces (e.g., *qacL* conferring resistance to quaternary ammonium disinfectants). As QACs are not ingested, genes conferring resistance to QACs likely originate from in-sewer selection as opposed to human gut microbiota (Maile-Moskowitz et al., 2025). Comparatively, *rsm*A and several beta-lactamases, were among the ARGs enriched in bulk wastewater (MaAslin3, p<0.05, **Figure 4c**). At the drug class level, aminoglycoside resistance genes—often associated with plasmids and integrons—were significantly enriched in biofilms (MaAslin3, p<0.05, **Figure 4d**). These patterns may reflect biofilms functioning as microbial reservoirs where repeated exposure to unmetabolized antibiotics, disinfectants, and other co-selecting stressors may promote AMR proliferation.^48,56^ Though current wastewater surveillance paradigms for AMR focus on shifts in prescription trends and resistant strains shaped by selection within the human microbiome, this framing does not account for in-sewer dynamics. Alternatively, our findings highlight that wastewater AMR signal is likely both from human shedding and environmental contributions. By identifying a distinct ‘pipe-resident’ resistome—characterized by biocide resistance (*qacL*) and opportunistic pathogen associations—we provide a basis for distinguishing environmental background signal from dynamic clinical inputs. This distinction is critical for preventing WS-derived overestimation of community AMR prevalence based on in-sewer proliferation.

#### Limitations

While this study identifies taxa, pathogens, and genes that overlap across sewer niches in a full-scale hospital setting, a key limitation is the assumption that shared taxa between wastewater and biofilms reflect biofilms acting as a microbial reservoir that seeds bulk wastewater. Alternatively, the reverse process (i.e., wastewater microorganisms depositing into biofilms) is possible, or pathogens could persist and grow in wastewater and biofilm independently. Bidirectional exchange of microorganisms and genetic material between these environments is likely;^48^ establishing directionality, however, is difficult in field-based observational studies. Controlled bench-scale experiments employing pseudo-tracer organisms introduced to each matrix would be better suited to quantify the direction and magnitude of microbial transfer between biofilms and bulk wastewater.

Though our research evaluates two major sources of microbial signal external to active hospital community shedding (biofilm and residual tap water contributions to wastewater pathogen and AMR signals), additional source contributions (e.g., plumbing sediments, in-sewer growth in bulk wastewater) were not directly within the scope of this study (i.e., a degree of sediments were likely captured by biofilm swabs). Given the short sewage travel times for building-scale wastewater surveillance prior to sample collection, bacterial growth within this window was assumed to be negligible.

While our metagenomic approach enabled broad detection of taxa and genes, other approaches could enhance taxonomic resolution, sensitivity, specificity, and contextualization of ARGs in bacterial hosts. For example, our taxonomic classification approach relied on read-based analysis due to only partial genome coverage of the probe-capture hybridization panel used, which may have reduced the specificity of pathogen and ARG assignments relative to assembly-based approaches. Comparison of sequenced isolates or metagenome-assembled genomes would enable more robust associations. Further, as with any total nucleic acid extraction and concentration protocol, methodological choices may introduce bias toward certain organisms. Sink drain and sewer biofilm samples were subjected to bead beating during extraction, whereas bulk wastewater samples that underwent probe-capture metagenomics were not. This difference in lysis efficiency could have enhanced detection of targets associated with more resilient cell structures in biofilms (e.g., mecA, Mycobacterium avium), potentially contributing to their absence in wastewater rather than reflecting true ecological differences.

#### Implications

Overall, our findings indicate that sewer biofilms represent a persistent and microbially diverse environment that could both reflect and reshape wastewater-derived signals. The high degree of overlap of ARGs and pathogens between biofilm and wastewater suggest that wastewater measurements integrate contributions from at least two ecologically distinct sources: dynamic human shedding inputs and pipe-resident microbial assemblages. Prior work has also suggested that wastewater and sewer biofilm communities — though distinct^57^— can substantially reflect infrastructure conditions and pipe-resident microbiomes, in both ARG and overall bacterial community composition.^31,57,58^ Further, as wastewater travel times and distance in-sewer increase, there is higher likelihood for interaction with biofilm, suggesting overlap in ARG and pathogen composition may be even more pronounced for wastewater collected at or near treatment facilities compared to the hospital-scale.^59^ Several clinically relevant opportunistic pathogens, including *Pseudomonas aeruginosa*, *Serratia marcescens* and *Klebsiella pneumoniae*, were consistently shared between biofilm and wastewater. These taxa likely represent persistent members of sewer biofilms that contribute continuously to wastewater signals. Patterns of antimicrobial resistance gene (ARG) enrichment further support the role of sewer biofilms as long-term reservoirs shaped by pipe-associated selection pressures, as ARGs enriched in biofilms were consistent with sustained exposure to disinfectants and other antimicrobials. As a result, wastewater resistome profiles may reflect both clinical inputs and in-sewer ecological processes, complicating direct inference of community-level antimicrobial resistance burden. Future bench-scale experiments could investigate the precise direction and magnitude of microbial movement between the biofilm and bulk wastewater to refine our understanding of these microbial reservoirs. Integrating baseline sewer infrastructure microbial ecology into surveillance models may thus be essential for improving both AMR monitoring and mechanistic understanding of pathogen dynamics in built water environments.

## Data Availability

All data produced in the present study are available upon reasonable request to the authors

https://github.com/avdarlingphd/darling-2026-biofilm-wbe

## Notes

### Competing Interest Statement

The authors have declared no competing interest.

