## Supplementary Information for "Biofilm Contributions of Bacterial Pathogens and Antimicrobial Resistance Genes to Wastewater Surveillance Signal at the Hospital Scale"

**Text**

**Genome Coverage Estimation for RPIP Panel Targets**

To validate Kraken2 classifications and estimate reference genome coverage for targeted pathogens, we developed a custom bioinformatics pipeline applied across the total 209 metagenomics samples processed with the Illumina Respiratory Pathogen Identification Panel (RPIP) hybridization-based probe capture enrichment protocol.

Reference genomes for RPIP panel targets detected in Kraken relative abundance output were downloaded from NCBI using the datasets command-line tool (v14). Of RPIP panel targets, 92 were detected across at least one sample and used for downstream analysis, yielding 5,269 unique sample-pathogen combinations. Reference genomes were organized by NCBI taxonomy ID and concatenated into single FASTA files where multiple chromosomes or genomic segments were present.

For each sample-pathogen combination, reads classified to the target taxon (including taxonomy below the species level) were extracted from Kraken2 output files using KrakenTools extract_kraken_reads.py with the --include-children flag. Extracted reads were aligned to the corresponding reference genome using BWA-MEM (v0.7.18) with 8 threads. Resulting alignments were coordinate-sorted and indexed using SAMtools (v1.17). Genome coverage metrics were calculated using mosdepth (v0.3.3), including mean depth, median depth, breadth of coverage at ≥1x, and breadth of coverage at ≥10x. For sample-pathogen combinations yielding no extracted reads, coverage metrics were recorded as zero without proceeding to alignment.

All 5,269 combinations were processed in parallel using a SLURM array job on the Harvard FASRC Cannon computing cluster. Results were aggregated into a single summary table for downstream analysis.

**BLAST validation**

BLASTN was used to validate Kraken2 taxonomic classifications. For each sample–taxid pair of interest, reads classified by Kraken2 to the target taxon were extracted using seqtk subseq and queried against the NCBI nucleotide (nt) database using BLASTN (task: blastn; threads: 8; max_target_seqs: 20; output format 6: qseqid, sscinames, pident, length, evalue, stitle). For each read, up to 20 BLAST hits were retained. Species names were taken from the sscinames field where available; otherwise, organism names were parsed from the stitle field by truncating at strain- or assembly-level descriptors (e.g., "strain", "subsp.", "chromosome", "complete genome"). A read was considered concordant with the Kraken2 classification if the expected organism appeared in **any** of its top 20 BLAST hits, rather than only the highest-scoring hit, to account for ties in alignment score across database entries. The fraction of concordant reads was averaged across R1 and R2. Sample–taxid pairs were classified as TRUE_POSITIVE if ≥80% of reads were concordant, FALSE_POSITIVE if <10% were concordant, and UNCERTAIN otherwise. Pairs for which no BLAST output was produced were labeled NO_DATA.

**Tables**

**Table S1.** Hospital Site Descriptions

| **Site** | **Patient Beds** | **Patient Toilets (+sink)** | **Patient Showers** | **Staff/public toilets (+ sink)** | **Handwash Sinks** | **Housekeeping Closets** | **Hoppers** | **Other** | **Units Captured** |
| --- | --- | --- | --- | --- | --- | --- | --- | --- | --- |
| Site 1 | 64 | 55 | 23 | 7 | 41 | 5 | 0 | peritoneal dialysis room, but seems to be without any drains, water treatment area | Ambulatory Surgery pre-op, Ambulatory Cardiology Procedure Recovery, Hemodialysis, Inpatient Internal Medicine, Ambulatory Research |
| Site 2 | 52 | 31 | 29 | 6 | 48 | 6 | 0 | 1 staff shower in call room | Trauma Surgery, Pediatric Neurology Cardiac ICU, Pediatric Sleep Lab |
| Site 3 | 78 | 49 | 24 | 7 | 28 | 5 | 0 | - | Hematology Oncology, General Medicine, Infectious Disease, Inpatient, General Surgery, General Medicine Dialysis, Inpatient Internal Medicine |
| Site 4 | 42 | 3 | 1 | 5 | 87 | 8 | 3 | - | Cardiac ICU, Surgical ICU, Surgery, Neurosurgery ICU |
| Site 5 | ~168 | ~22+ | - | 4 | 3 | 4 | 4 | 1 scope washer, 9 patient bathrooms were staff/patient | Radiation Therapy, Observation, Radiology, OR/procedure, Infusion, General Clinical |

**Table S2.** Methodological issues for 16S sequencing and hybrid metagenomic samples.

| Location | Type | Timepoint | Issue | Count |
| --- | --- | --- | --- | --- |
| Site 5 | Wastewater | 1 | Below 24 Hours | 1  9/5/2024 |
| Site 3 | Wastewater | 1 | Below 24 Hours | 1  9/5/2024 |
| Site 1 | Wastewater | 1 | Below 24 Hours | 1  9/5/2024 |
| Site 2 | Wastewater | 1 | Below 24 Hours | 1  9/5/2024 |
| Site 4 | Wastewater | 1 | Below 24 Hours | 1  9/5/2024 |
| Site 4 | Wastewater | 1 | Grab Sample | 2  9/12/2024  9/30/2024 |
| Site 5 | Wastewater | 1 | Grab Sample | 1  9/25/2024 |
| Site 5 | Sink Drain | 2 | Failed Metagenomic Sequencing | 2  12/11/2024  12/11/2024 |
| Site 5 | Wastewater | 2 | Grab Sample | 4  10/10/2024  10/17/2024  10/24/2024  11/14/2024 |
| Site 4 | Wastewater | 2 | Grab Sample | 3  10/3/2024  10/7/2024  10/14/2024 |
| Site 2 | Wastewater | 2 | Grab Sample | 1  10/28/2024 |
| Site 1 | Wastewater | 2 | Grab Sample | 1  11/7/2024 |
| Site 2 | Branch Drain | 2 | Failed 16s Sequencing | 1  10/28/2024 |
| Site 1 | Wastewater | 3 | Grab Sample | 1  12/5/2024 |

**Table S3.** Sample collection timeline

|  | **Dec (2023) - Apr (2024)** | | **Aug - Sep (2024)** | | **Oct - Nov (2024)** | | **Dec (2024)** | |
| --- | --- | --- | --- | --- | --- | --- | --- | --- |
|  | **Pre-Study Collection** | | **Timepoint 1** | | **Timepoint 2** | | **Timepoint 3** | |
|  | **16S** | **RPIP MGX** | **16S** | **RPIP MGX** | **16S** | **RPIP MGX** | **16S** | **RPIP MGX** |
| **Tap** | 29 | 0 | 0 | 0 | 0 | 0 | 0 | 0 |
| **Sink Biofilm** | 0 | 0 | 15 | 0 | 15 | 0 | 15 | 13 |
| **Sewer Biofilm** | 0 | 0 | 5 | 5 | 5 | 5 | 5 | 5 |
| **Waste-water** | 0 | 0 | 10 | 52 | 10 | 110 | 10 | 19 |
| *Notes:* RPIP MGX: hybridization-based probe-capture metagenomic samples | | | | | | | | |

**Table S4.** Respiratory Pathogen ID/AMR (RPIP) panel targets

| **Organism Type** | **Species** |
| --- | --- |
| **Bacteria** | *Achromobacter denitrificans; Achromobacter xylosoxidans; Acinetobacter baumannii; Acinetobacter lwoffii; Acinetobacter nosocomialis; Acinetobacter pittii; Actinomyces graevenitzii; Actinomyces israelii; Actinomyces naeslundii; Aeromonas caviae; Aeromonas hydrophila; Aeromonas sobria; Aeromonas veronii; Aggregatibacter actinomycetemcomitans; Aggregatibacter aphrophilus; Arcanobacterium haemolyticum; Bacillus anthracis; Bacillus cereus; Bacillus thuringiensis; Bacteroides fragilis; Bartonella henselae; Bartonella quintana; Bordetella bronchiseptica; Bordetella hinzii; Bordetella holmesii; Bordetella parapertussis; Bordetella pertussis; Bordetella petrii; Brucella abortus; Brucella canis; Brucella melitensis; Brucella suis; Burkholderia cepacia complex; Burkholderia gladioli; Burkholderia glumae; Burkholderia mallei; Burkholderia pseudomallei; Burkholderia thailandensis; Campylobacter concisus; Capnocytophaga gingivalis; Capnocytophaga leadbetteri; Capnocytophaga sputigena; Cardiobacterium hominis; Cardiobacterium valvarum; Chlamydia pneumoniae; Chlamydia psittaci; Chlamydia trachomatis; Chromobacterium violaceum; Citrobacter freundii complex; Citrobacter koseri; Corynebacterium diphtheriae; Corynebacterium jeikeium; Corynebacterium propinquum; Corynebacterium pseudodiphtheriticum; Corynebacterium pseudotuberculosis; Corynebacterium striatum; Corynebacterium ulcerans; Coxiella burnetii; Cronobacter sakazakii; Delftia acidovorans; Dialister pneumosintes; Dolosigranulum pigrum; Eikenella corrodens; Elizabethkingia anophelis; Elizabethkingia meningoseptica; Enterobacter cloacae complex; Enterococcus faecalis; Enterococcus faecium; Escherichia coli; Eubacterium brachy; Eubacterium limosum; Eubacterium nodatum; Finegoldia magna; Francisella tularensis; Fusobacterium necrophorum; Fusobacterium nucleatum; Gemella haemolysans; Gemella morbillorum; Gordonia araii; Gordonia bronchialis; Haemophilus haemolyticus; Haemophilus influenzae; Haemophilus parahaemolyticus; Haemophilus parainfluenzae; Haemophilus pittmaniae; Hafnia alvei; Kingella kingae; Klebsiella aerogenes; Klebsiella oxytoca; Klebsiella pneumoniae; Klebsiella quasipneumoniae; Klebsiella variicola; Kytococcus sedentarius; Leclercia adecarboxylata; Legionella anisa; Legionella feeleii; Legionella longbeachae; Legionella pneumophila; Legionella wadsworthii; Leptospira interrogans; Leptotrichia buccalis; Listeria monocytogenes; Moraxella catarrhalis; Moraxella osloensis; Morganella morganii; Mycobacterium avium complex; Mycobacterium gordonae; Mycobacterium kansasii; Mycobacterium malmoense; Mycobacterium scrofulaceum; Mycobacterium simiae complex; Mycobacterium szulgai; Mycobacterium tuberculosis complex; Mycobacterium xenopi; Mycolicibacterium fortuitum; Mycobacteroides abscessus; Mycobacteroides chelonae; Mycobacteroides immunogenum; Mycoplasma pneumoniae; Neisseria flavescens; Neisseria lactamica; Neisseria meningitidis; Neisseria mucosa; Nocardia abscessus; Nocardia arthritidis; Nocardia beijingensis; Nocardia brasiliensis; Nocardia cyriacigeorgica; Nocardia farcinica; Nocardia nova; Nocardia otitidiscaviarum; Nocardia transvalensis; Nocardia veterana; Ochrobactrum anthropi; Orientia tsutsugamushi; Pandoraea pulmonicola; Pantoea agglomerans; Parvimonas micra; Pasteurella multocida; Pediococcus acidilactici; Peptostreptococcus anaerobius; Prevotella buccae; Prevotella intermedia; Prevotella melaninogenica; Prevotella pleuritidis; Proteus mirabilis; Proteus penneri; Proteus vulgaris; Providencia stuartii; Pseudomonas aeruginosa; Pseudomonas fluorescens; Pseudomonas stutzeri; Ralstonia pickettii; Raoultella ornithinolytica; Raoultella planticola; Rhodococcus hoagii; Rickettsia rickettsii; Rothia mucilaginosa; Salmonella enterica; Serratia marcescens; Shewanella putrefaciens; Slackia exigua; Sphingomonas paucimobilis; Staphylococcus aureus; Stenotrophomonas maltophilia; Streptococcus agalactiae; Streptococcus anginosus; Streptococcus constellatus; Streptococcus dysgalactiae; Streptococcus intermedius; Streptococcus mitis; Streptococcus pneumoniae; Streptococcus pyogenes; Tatlockia micdadei; Treponema denticola; Tropheryma whipplei; Tsukamurella pulmonis; Tsukamurella tyrosinosolvens; Ureaplasma parvum; Ureaplasma urealyticum; Veillonella parvula; Williamsia muralis; Yersinia enterocolitica; Yersinia pestis* |
| **Viruses** | Coxsackievirus A; Coxsackievirus B; Cytomegalovirus (CMV); Enterovirus A71; Enterovirus D68; Epstein-Barr virus (EBV); Herpes simplex virus 1 (HSV-1); Human adenovirus B; Human adenovirus C; Human adenovirus E; Human bocavirus 1; Human coronavirus 229E; Human coronavirus HKU1; Human coronavirus NL63; Human coronavirus OC43; Human herpesvirus 6 (HHV-6); Human metapneumovirus; Human parainfluenza virus 1; Human parainfluenza virus 2; Human parainfluenza virus 3; Human parainfluenza virus 4; Human parechovirus; Human rhinovirus A; Human rhinovirus B; Human rhinovirus C; Influenza A virus (H1N1); Influenza A virus (H3N2); Influenza A virus (H5N1); Influenza A virus (H7N9); Influenza A virus (H9N2); Influenza B virus; Influenza C virus; MERS coronavirus (MERS-CoV); Measles virus; Mumps virus; Parvovirus B19; Respiratory syncytial virus A; Respiratory syncytial virus B; Rubella virus; SARS coronavirus; SARS-CoV-2 (2019-nCoV); Varicella-zoster virus (VZV) |
| **Fungi** | *Alternaria alternata; Alternaria infectoria; Apophysomyces elegans; Aspergillus flavus; Aspergillus fumigatus; Aspergillus nidulans; Aspergillus niger; Aspergillus terreus; Aspergillus versicolor; Blastomyces dermatitidis; Byssochlamys spectabilis; Candida auris; Cladophialophora bantiana; Coccidioides immitis; Coccidioides posadasii; Cryptococcus neoformans/Cryptococcus gattii; Cunninghamella bertholletiae; Curvularia geniculata; Curvularia lunata; Exophiala dermatitidis; Fusarium oxysporum; Fusarium proliferatum; Fusarium solani; Fusarium verticillioides; Histoplasma capsulatum; Lichtheimia corymbifera; Lichtheimia ramosa; Lomentospora prolificans; Microascus cinereus; Microascus cirrosus; Microascus paisii; Mucor circinelloides; Mucor indicus; Mucor racemosus; Paracoccidioides brasiliensis; Pneumocystis jirovecii; Purpureocillium lilacinum; Rasamsonia aegroticola; Rasamsonia argillacea; Rhizomucor pusillus; Rhizopus azygosporus; Rhizopus microsporus; Rhizopus oryzae; Saksenaea vasiformis; Sarocladium kiliense; Scedosporium apiospermum; Schizophyllum commune; Scopulariopsis brevicaulis; Sporothrix schenckii; Syncephalastrum racemosum; Talaromyces marneffei; Trichosporon asahii* |

**Table S5.** 16S Sequencing Successfully Sequenced Samples.

|  | Sample Type | | | |
| --- | --- | --- | --- | --- |
|  | **Tap** | **Drain** | **Branch Drain** | **Wastewater** |
| Site 1 | 0 | Sep: 3  Oct: 3  Dec: 3 | Sep: 1  Oct: 1  Dec: 1 | Sep: 2  Oct: 2  Dec: 2 |
| Site 3 | 0 | Sep: 3  Oct: 3  Dec: 3 | Sep: 1  Oct: 1  Dec: 1 | Sep: 2  Oct: 2  Dec: 2 |
| Site 4 | 0 | Sep: 3  Oct: 3  Dec: 3 | Sep: 1  Oct: 1  Dec: 1 | Sep: 2  Oct: 2  Dec: 2 |
| Site 2 | 0 | Sep: 3  Oct: 3  Dec: 3 | Sep: 1  Oct: 0  Dec: 1 | Sep: 2  Oct: 2  Dec: 2 |
| Site 5 | 36 | Sep: 3  Oct: 3  Dec: 3 | Sep: 1  Oct: 1  Dec: 1 | Sep: 2  Oct: 2  Dec: 2 |

**Table S6.** Total Reads QC’d, Trimmed, and Dehosted and Total Reads Annotated by Kraken2

| **Date** | **Location** | **Type** | **Total Reads Annotated by Kraken** | **Total Reads After Trimming, Dehosting, and QC** | **Timepoint** |
| --- | --- | --- | --- | --- | --- |
| 2024-10-02 | Site 4 | Wastewater | 476370 | 43117400 | 2 |
| 2024-10-02 | Site 2 | Wastewater | 158549 | 22538007 | 2 |
| 2024-10-02 | Site 5 | Wastewater | 650325 | 23375398 | 2 |
| 2024-11-13 | Site 4 | Wastewater | 461013 | 21188967 | 2 |
| 2024-11-13 | Site 2 | Wastewater | 570289 | 14556712 | 2 |
| 2024-11-13 | Site 1 | Wastewater | 326591 | 54491707 | 2 |
| 2024-11-13 | Site 3 | Wastewater | 423334 | 13438650 | 2 |
| 2024-11-13 | Site 5 | Wastewater | 642301 | 14847871 | 2 |
| 2024-09-05 | Site 5 | Wastewater | 753158 | 26726396 | 1 |
| 2024-09-05 | Site 3 | Wastewater | 743697 | 28214946 | 1 |
| 2024-09-05 | Site 1 | Wastewater | 543311 | 20732220 | 1 |
| 2024-09-05 | Site 2 | Wastewater | 632868 | 49983231 | 1 |
| 2024-09-05 | Site 4 | Wastewater | 979627 | 14868274 | 1 |
| 2024-09-09 | Site 3 | Wastewater | 413500 | 32136905 | 1 |
| 2024-09-09 | Site 4 | Wastewater | 281557 | 47280917 | 1 |
| 2024-09-09 | Site 5 | Wastewater | 724245 | 17258322 | 1 |
| 2024-09-11 | Site 5 | Wastewater | 573153 | 10372498 | 1 |
| 2024-09-11 | Site 4 | Wastewater | 716463 | 19572869 | 1 |
| 2024-09-11 | Site 3 | Wastewater | 279850 | 9899594 | 1 |
| 2024-09-11 | Site 2 | Wastewater | 453487 | 30808071 | 1 |
| 2024-09-11 | Site 1 | Wastewater | 495876 | 24830232 | 1 |
| 2024-09-16 | Site 3 | Wastewater | 976441 | 51019943 | 1 |
| 2024-09-16 | Site 2 | Wastewater | 326505 | 37616605 | 1 |
| 2024-09-16 | Site 5 | Wastewater | 601024 | 16138761 | 1 |
| 2024-09-16 | Site 4 | Wastewater | 444948 | 23550694 | 1 |
| 2024-09-16 | Site 1 | Wastewater | 331541 | 25981064 | 1 |
| 2024-09-12 | Site 3 | Wastewater | 523034 | 18952264 | 1 |
| 2024-09-12 | Site 1 | Wastewater | 476967 | 42014011 | 1 |
| 2024-09-12 | Site 4 | Wastewater | 425397 | 37838995 | 1 |
| 2024-09-12 | Site 2 | Wastewater | 662889 | 37194773 | 1 |
| 2024-10-09 | Site 2 | Wastewater | 433086 | 39621763 | 2 |
| 2024-09-18 | Site 3 | Wastewater | 604693 | 16877457 | 1 |
| 2024-09-18 | Site 4 | Wastewater | 466758 | 28786519 | 1 |
| 2024-09-18 | Site 2 | Wastewater | 284977 | 41596159 | 1 |
| 2024-09-18 | Site 5 | Wastewater | 1360057 | 16704593 | 1 |
| 2024-09-18 | Site 1 | Wastewater | 252392 | 31111627 | 1 |
| 2024-09-19 | Site 3 | Wastewater | 678327 | 17363405 | 1 |
| 2024-09-19 | Site 5 | Wastewater | 822079 | 39119121 | 1 |
| 2024-09-19 | Site 1 | Wastewater | 376865 | 36747451 | 1 |
| 2024-09-19 | Site 2 | Wastewater | 339530 | 44926939 | 1 |
| 2024-09-19 | Site 4 | Wastewater | 355661 | 16622240 | 1 |
| 2024-09-23 | Site 2 | Wastewater | 387899 | 44329608 | 1 |
| 2024-09-23 | Site 4 | Wastewater | 505443 | 52970334 | 1 |
| 2024-09-23 | Site 5 | Wastewater | 420016 | 10178984 | 1 |
| 2024-09-23 | Site 3 | Wastewater | 382059 | 10461256 | 1 |
| 2024-09-23 | Site 1 | Wastewater | 262592 | 15866586 | 1 |
| 2024-09-25 | Site 5 | Wastewater | 98584 | 8576601 | 1 |
| 2024-09-25 | Site 1 | Wastewater | 70604 | 7661797 | 1 |
| 2024-09-25 | Site 3 | Wastewater | 84450 | 2766393 | 1 |
| 2024-09-25 | Site 4 | Wastewater | 538131 | 17704276 | 1 |
| 2024-09-25 | Site 2 | Wastewater | 446561 | 38680587 | 1 |
| 2024-10-07 | Site 5 | Wastewater | 635018 | 16449640 | 2 |
| 2024-10-28 | Site 1 | Wastewater | 857423 | 39415680 | 2 |
| 2024-11-04 | Site 1 | Wastewater | 289532 | 40598822 | 2 |
| 2024-11-18 | Site 1 | Wastewater | 195155 | 25300983 | 2 |
| 2024-11-18 | Site 3 | Wastewater | 295271 | 8175791 | 2 |
| 2024-11-18 | Site 5 | Wastewater | 618878 | 22401981 | 2 |
| 2024-11-18 | Site 2 | Wastewater | 292690 | 53689063 | 2 |
| 2024-11-18 | Site 4 | Wastewater | 770531 | 42769123 | 2 |
| 2024-10-10 | Site 2 | Wastewater | 544592 | 40371783 | 2 |
| 2024-10-10 | Site 4 | Wastewater | 260845 | 41406786 | 2 |
| 2024-10-10 | Site 3 | Wastewater | 689069 | 16382711 | 2 |
| 2024-10-10 | Site 1 | Wastewater | 461962 | 38275049 | 2 |
| 2024-10-10 | Site 5 | Wastewater | 750215 | 13490872 | 2 |
| 2024-10-03 | Site 1 | Wastewater | 384235 | 36190922 | 2 |
| 2024-10-03 | Site 3 | Wastewater | 371078 | 26607422 | 2 |
| 2024-10-03 | Site 5 | Wastewater | 506053 | 20165888 | 2 |
| 2024-10-03 | Site 2 | Wastewater | 260201 | 24312836 | 2 |
| 2024-10-03 | Site 4 | Wastewater | 301361 | 35913712 | 2 |
| 2024-12-05 | Site 4 | Wastewater | 826315 | 36619573 | 3 |
| 2024-12-05 | Site 3 | Wastewater | 498436 | 13969774 | 3 |
| 2024-12-05 | Site 5 | Wastewater | 547843 | 18300009 | 3 |
| 2024-12-05 | Site 1 | Wastewater | 363327 | 63912293 | 3 |
| 2024-10-09 | Site 5 | Wastewater | 595110 | 16665180 | 2 |
| 2024-10-09 | Site 4 | Wastewater | 485115 | 45132250 | 2 |
| 2024-10-09 | Site 3 | Wastewater | 790260 | 17769266 | 2 |
| 2024-10-09 | Site 1 | Wastewater | 271769 | 18541634 | 2 |
| 2024-10-17 | Site 5 | Wastewater | 565390 | 18123475 | 2 |
| 2024-10-17 | Site 2 | Wastewater | 383793 | 35125684 | 2 |
| 2024-10-17 | Site 4 | Wastewater | 467678 | 34601177 | 2 |
| 2024-10-17 | Site 3 | Wastewater | 593735 | 9811716 | 2 |
| 2024-10-17 | Site 1 | Wastewater | 622076 | 64114238 | 2 |
| 2024-10-24 | Site 4 | Wastewater | 574143 | 9647328 | 2 |
| 2024-10-24 | Site 2 | Wastewater | 466071 | 50440559 | 2 |
| 2024-10-24 | Site 3 | Wastewater | 616200 | 25744320 | 2 |
| 2024-10-24 | Site 1 | Wastewater | 324295 | 30720286 | 2 |
| 2024-10-24 | Site 5 | Wastewater | 1001783 | 15724887 | 2 |
| 2024-10-30 | Site 4 | Wastewater | 1380687 | 26816400 | 2 |
| 2024-10-30 | Site 1 | Wastewater | 529014 | 41094846 | 2 |
| 2024-10-30 | Site 3 | Wastewater | 504902 | 14597609 | 2 |
| 2024-10-30 | Site 5 | Wastewater | 585732 | 23367783 | 2 |
| 2024-11-25 | Site 2 | Wastewater | 172971 | 42563059 | 2 |
| 2024-11-25 | Site 4 | Wastewater | 373701 | 16535354 | 2 |
| 2024-11-25 | Site 5 | Wastewater | 596101 | 17749187 | 2 |
| 2024-11-25 | Site 3 | Wastewater | 581334 | 14847275 | 2 |
| 2024-11-25 | Site 1 | Wastewater | 330711 | 23597601 | 2 |
| 2024-11-21 | Site 2 | Wastewater | 163299 | 43537140 | 2 |
| 2024-11-21 | Site 3 | Wastewater | 412544 | 23249262 | 2 |
| 2024-11-21 | Site 1 | Wastewater | 636513 | 46037937 | 2 |
| 2024-11-21 | Site 4 | Wastewater | 477589 | 22698885 | 2 |
| 2024-12-02 | Site 1 | Wastewater | 438584 | 37176870 | 3 |
| 2024-12-02 | Site 3 | Wastewater | 531200 | 33244505 | 3 |
| 2024-12-02 | Site 5 | Wastewater | 632999 | 17755704 | 3 |
| 2024-12-02 | Site 2 | Wastewater | 232664 | 28036353 | 3 |
| 2024-12-02 | Site 4 | Wastewater | 583143 | 31715532 | 3 |
| 2024-11-14 | Site 3 | Wastewater | 195943 | 21475767 | 2 |
| 2024-11-14 | Site 5 | Wastewater | 432076 | 15353491 | 2 |
| 2024-11-14 | Site 2 | Wastewater | 1224540 | 38463511 | 2 |
| 2024-11-14 | Site 1 | Wastewater | 443261 | 57767564 | 2 |
| 2024-11-11 | Site 1 | Wastewater | 271957 | 18681057 | 2 |
| 2024-11-20 | Site 4 | Wastewater | 802644 | 22623723 | 2 |
| 2024-11-20 | Site 1 | Wastewater | 282593 | 21633530 | 2 |
| 2024-11-20 | Site 3 | Wastewater | 649470 | 30946465 | 2 |
| 2024-10-31 | Site 2 | Wastewater | 579520 | 23291898 | 2 |
| 2024-10-31 | Site 4 | Wastewater | 399899 | 18953885 | 2 |
| 2024-10-31 | Site 3 | Wastewater | 100532 | 7997384 | 2 |
| 2024-10-31 | Site 5 | Wastewater | 1057260 | 90385838 | 2 |
| 2024-11-06 | Site 2 | Wastewater | 276494 | 9473793 | 2 |
| 2024-11-06 | Site 4 | Wastewater | 865113 | 24610882 | 2 |
| 2024-11-06 | Site 1 | Wastewater | 959838 | 42928459 | 2 |
| 2024-11-06 | Site 3 | Wastewater | 701546 | 24117144 | 2 |
| 2024-11-06 | Site 5 | Wastewater | 1023697 | 32823015 | 2 |
| 2024-10-30 | Site 2 | Wastewater | 749157 | 36995864 | 2 |
| 2024-11-04 | Site 2 | Wastewater | 390525 | 17579344 | 2 |
| 2024-11-04 | Site 4 | Wastewater | 427859 | 16836708 | 2 |
| 2024-11-04 | Site 5 | Wastewater | 1458441 | 48577659 | 2 |
| 2024-11-04 | Site 3 | Wastewater | 238834 | 8190769 | 2 |
| 2024-12-04 | Site 1 | Wastewater | 481765 | 25714228 | 3 |
| 2024-12-04 | Site 3 | Wastewater | 985430 | 17148401 | 3 |
| 2024-12-04 | Site 5 | Wastewater | 408574 | 12122954 | 3 |
| 2024-12-04 | Site 2 | Wastewater | 760368 | 11144722 | 3 |
| 2024-12-04 | Site 4 | Wastewater | 1244934 | 22040287 | 3 |
| 2024-12-11 | Site 2 | Wastewater | 574145 | 49367626 | 3 |
| 2024-12-11 | Site 4 | Wastewater | 252165 | 8343034 | 3 |
| 2024-12-11 | Site 5 | Wastewater | 850165 | 13513706 | 3 |
| 2024-12-11 | Site 3 | Wastewater | 359649 | 21773999 | 3 |
| 2024-12-09 | Site 1 | Wastewater | 605094 | 68014686 | 3 |
| 2024-10-28 | Site 3 | Wastewater | 744275 | 25637304 | 2 |
| 2024-10-28 | Site 5 | Wastewater | 219370 | 13815374 | 2 |
| 2024-10-28 | Site 4 | Wastewater | 1354067 | 20767320 | 2 |
| 2024-10-28 | Site 2 | Wastewater | 238057 | 32616277 | 2 |
| 2024-10-21 | Site 5 | Wastewater | 660279 | 48404071 | 2 |
| 2024-10-21 | Site 4 | Wastewater | 492453 | 17877116 | 2 |
| 2024-10-21 | Site 2 | Wastewater | 1182883 | 19286621 | 2 |
| 2024-10-16 | Site 3 | Wastewater | 735246 | 25161998 | 2 |
| 2024-10-23 | Site 3 | Wastewater | 732307 | 19968274 | 2 |
| 2024-10-23 | Site 5 | Wastewater | 119867 | 27941805 | 2 |
| 2024-10-23 | Site 2 | Wastewater | 145909 | 37064000 | 2 |
| 2024-10-23 | Site 4 | Wastewater | 726840 | 18983928 | 2 |
| 2024-10-23 | Site 1 | Wastewater | 431196 | 32554636 | 2 |
| 2024-10-16 | Site 4 | Wastewater | 490931 | 57653771 | 2 |
| 2024-10-16 | Site 2 | Wastewater | 337489 | 16739702 | 2 |
| 2024-09-26 | Site 3 | Wastewater | 759268 | 32468396 | 1 |
| 2024-11-07 | Site 1 | Wastewater | 486688 | 59410898 | 2 |
| 2024-09-25 | Site 5 | Wastewater | 613404 | 12215107 | 1 |
| 2024-11-07 | Site 5 | Wastewater | 392634 | 21629363 | 2 |
| 2024-11-07 | Site 3 | Wastewater | 279865 | 15853694 | 2 |
| 2024-10-14 | Site 1 | Wastewater | 481475 | 57617017 | 2 |
| 2024-11-07 | Site 2 | Wastewater | 398558 | 39323689 | 2 |
| 2024-10-14 | Site 3 | Wastewater | 628630 | 9135445 | 2 |
| 2024-10-14 | Site 4 | Wastewater | 340419 | 43008130 | 2 |
| 2024-10-14 | Site 2 | Wastewater | 343383 | 14798967 | 2 |
| 2024-10-16 | Site 1 | Wastewater | 439028 | 43497616 | 2 |
| 2024-10-14 | Site 5 | Wastewater | 621865 | 16394908 | 2 |
| 2024-09-26 | Site 1 | Wastewater | 396906 | 41344214 | 1 |
| 2024-09-26 | Site 2 | Wastewater | 176002 | 10438470 | 1 |
| 2024-09-26 | Site 4 | Wastewater | 469080 | 63598163 | 1 |
| 2024-09-30 | Site 1 | Wastewater | 428251 | 18330646 | 1 |
| 2024-09-30 | Site 2 | Wastewater | 437975 | 28650582 | 1 |
| 2024-09-30 | Site 5 | Wastewater | 231460 | 10675620 | 1 |
| 2024-09-30 | Site 4 | Wastewater | 458334 | 17631216 | 1 |
| 2024-09-30 | Site 3 | Wastewater | 355665 | 15227207 | 1 |
| 2024-11-11 | Site 3 | Wastewater | 171124 | 22060725 | 2 |
| 2024-11-11 | Site 5 | Wastewater | 624435 | 21914026 | 2 |
| 2024-11-11 | Site 4 | Wastewater | 634177 | 36680005 | 2 |
| 2024-11-11 | Site 2 | Wastewater | 390894 | 11614498 | 2 |
| 2024-10-02 | Site 1 | Wastewater | 493309 | 31187158 | 2 |
| 2024-10-07 | Site 1 | Wastewater | 342166 | 25593997 | 2 |
| 2024-10-07 | Site 3 | Wastewater | 676118 | 19121332 | 2 |
| 2024-10-07 | Site 4 | Wastewater | 518633 | 61181434 | 2 |
| 2024-10-07 | Site 2 | Wastewater | 470162 | 10068587 | 2 |
| 2024-08-27 | Site 3 | Sewer Biofilm | 588082 | 37138413 | 1 |
| 2024-08-29 | Site 2 | Sewer Biofilm | 178603 | 17976866 | 1 |
| 2024-08-27 | Site 1 | Sewer Biofilm | 348256 | 35620253 | 1 |
| 2024-12-11 | Site 4 | Sink Biofilm | 348271 | 46816007 | 3 |
| 2024-12-11 | Site 2 | Sink Biofilm | 767779 | 33514425 | 3 |
| 2024-12-11 | Site 2 | Sink Biofilm | 487080 | 24393455 | 3 |
| 2024-12-11 | Site 2 | Sink Biofilm | 797515 | 32344105 | 3 |
| 2024-12-11 | Site 4 | Sink Biofilm | 1696922 | 1.92E+08 | 3 |
| 2024-12-11 | Site 4 | Sink Biofilm | 2785699 | 1.08E+08 | 3 |
| 2024-12-12 | Site 3 | Sewer Biofilm | 834918 | 47385329 | 3 |
| 2024-12-12 | Site 5 | Sewer Biofilm | 312227 | 24499455 | 3 |
| 2024-12-11 | Site 3 | Sink Biofilm | 306426 | 23612678 | 3 |
| 2024-12-11 | Site 1 | Sink Biofilm | 211676 | 21914700 | 3 |
| 2024-12-11 | Site 3 | Sink Biofilm | 384413 | 45841654 | 3 |
| 2024-12-11 | Site 1 | Sink Biofilm | 775329 | 36874377 | 3 |
| 2024-12-11 | Site 1 | Sink Biofilm | 1064315 | 26879285 | 3 |
| 2024-12-11 | Site 3 | Sink Biofilm | 370298 | 23604642 | 3 |
| 2024-12-11 | Site 5 | Sink Biofilm | 411461 | 34877918 | 3 |
| 2024-08-29 | Site 4 | Sewer Biofilm | 457352 | 34582024 | 1 |
| 2024-08-30 | Site 5 | Sewer Biofilm | 499089 | 29793482 | 1 |
| 2024-10-28 | Site 4 | Sewer Biofilm | 449470 | 35242462 | 2 |
| 2024-10-28 | Site 2 | Sewer Biofilm | 896651 | 44100374 | 2 |
| 2024-10-28 | Site 1 | Sewer Biofilm | 335452 | 20421058 | 2 |
| 2024-10-28 | Site 3 | Sewer Biofilm | 466032 | 23789089 | 2 |
| 2024-10-28 | Site 5 | Sewer Biofilm | 849974 | 54973986 | 2 |
| 2024-12-12 | Site 4 | Sewer Biofilm | 613347 | 15000558 | 3 |
| 2024-12-12 | Site 2 | Sewer Biofilm | 627844 | 38414554 | 3 |
| 2024-12-12 | Site 1 | Sewer Biofilm | 688806 | 52224964 | 3 |

**Table S7.** 16S rRNA gene amplicon sequencing-derived PERMANOVA

|  | R2 | F | p-value |
| --- | --- | --- | --- |
| Model | 0.286 | 15.221 | <0.001 |
| Residual | 0.714 |  |  |
| Total | 1 |  |  |

**Table S8.** 16S rRNA gene amplicon sequencing-derived pairwise PERMANOVA

| Pairs | R2 | F | Adjusted p-value |
| --- | --- | --- | --- |
| Tap Water vs Sink Biofilm | 0.232 | 21.697 | <0.006 |
| Tap Water vs Sewer Biofilm | 0.333 | 20.423 | <0.006 |
| Tap Water vs Wastewater | 0.320 | 26.774 | <0.006 |
| Sink Biofilm vs Sewer Biofilm | 0.128 | 8.386 | <0.006 |
| Sink Biofilm vs Wastewater | 0.163 | 14.295 | <0.006 |
| Sewer Biofilm vs Wastewater | 0.054 | 2.4144 | <0.006 |

**Table S9.** Probe-based Metagenomic sequencing-derived pairwise PERMANOVA

| Pairs | R2 | F | Adjusted p-value |
| --- | --- | --- | --- |
| Sink Biofilm vs Sewer Biofilm | 0.33 | 12.7 | <0.003 |
| Sink Biofilm vs Wastewater | 0.32 | 12.1 | <0.003 |
| Sewer Biofilm vs Wastewater | 0.06 | 1.92 | 0.303 |

**Table S10.** Probe-based Metagenomic sequencing-derived Pairwise PERMANOVA on Resistome

| Pairs | R2 | F | Adjusted p-value |
| --- | --- | --- | --- |
| Sink Biofilm vs Sewer Biofilm | 0.375 | 15.58 | <0.003 |
| Sink Biofilm vs Wastewater | 0.431 | 19.73 | <0.003 |
| Sewer Biofilm vs Wastewater | 0.283 | 11.03 | <0.003 |

**Figures**

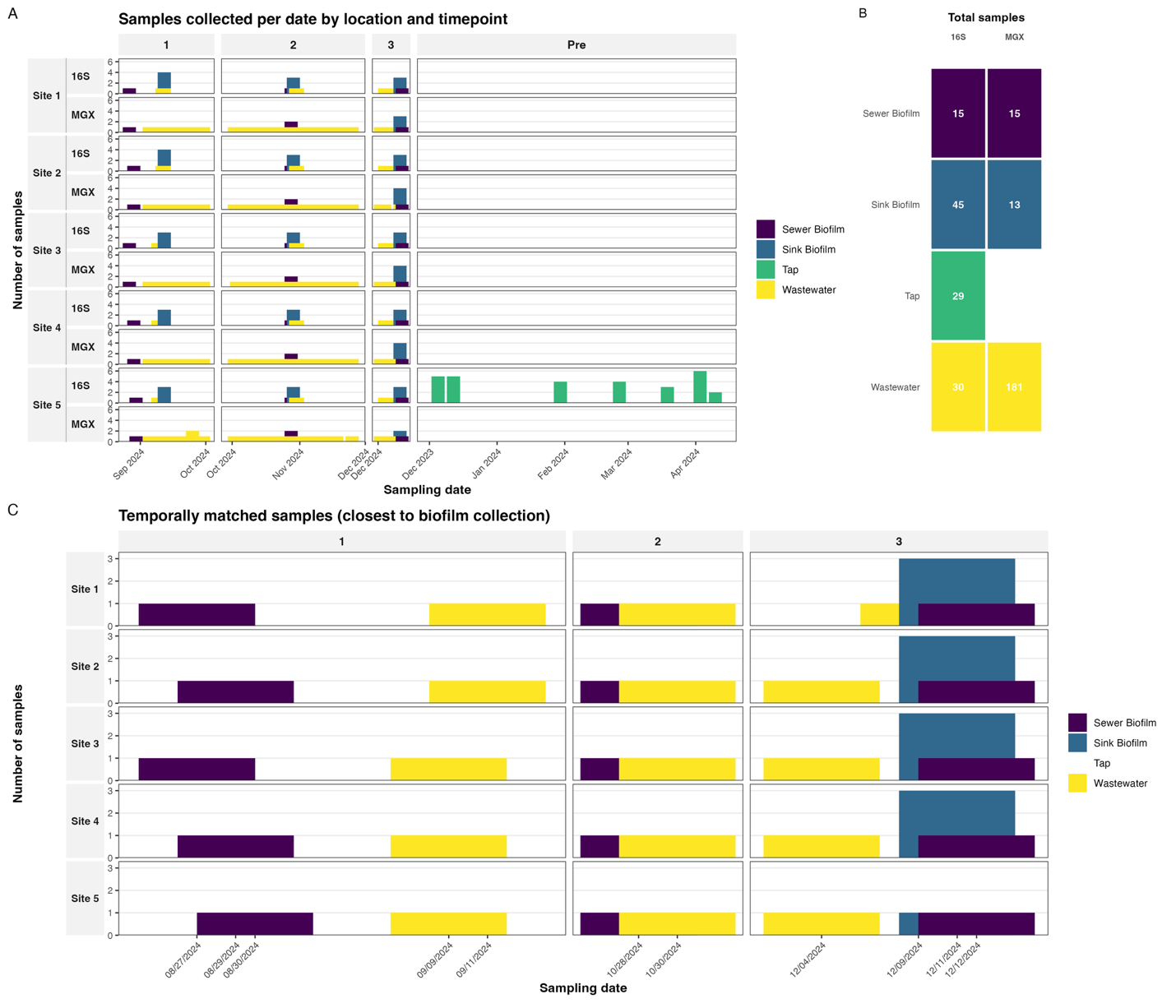

**Figure S1.** **Sample collection dates for each sample type and sequencing approach**. (a) Number of samples collected per date at each site, stratified by sequencing method and grouped by timepoint (Pre, 1, 2, 3). Each bar represents the count of samples collected on a given date, colored by sample type. (b) Total sample counts aggregated across all sites and timepoints, broken down by sequencing method and sample type. (c) Temporally matched subset of MGX samples retained for paired analyses with sewer biofilm collections. For each site × timepoint, sewer biofilm sampling dates served as the reference, and the nearest-in-time wastewater and sink biofilm samples were selected.

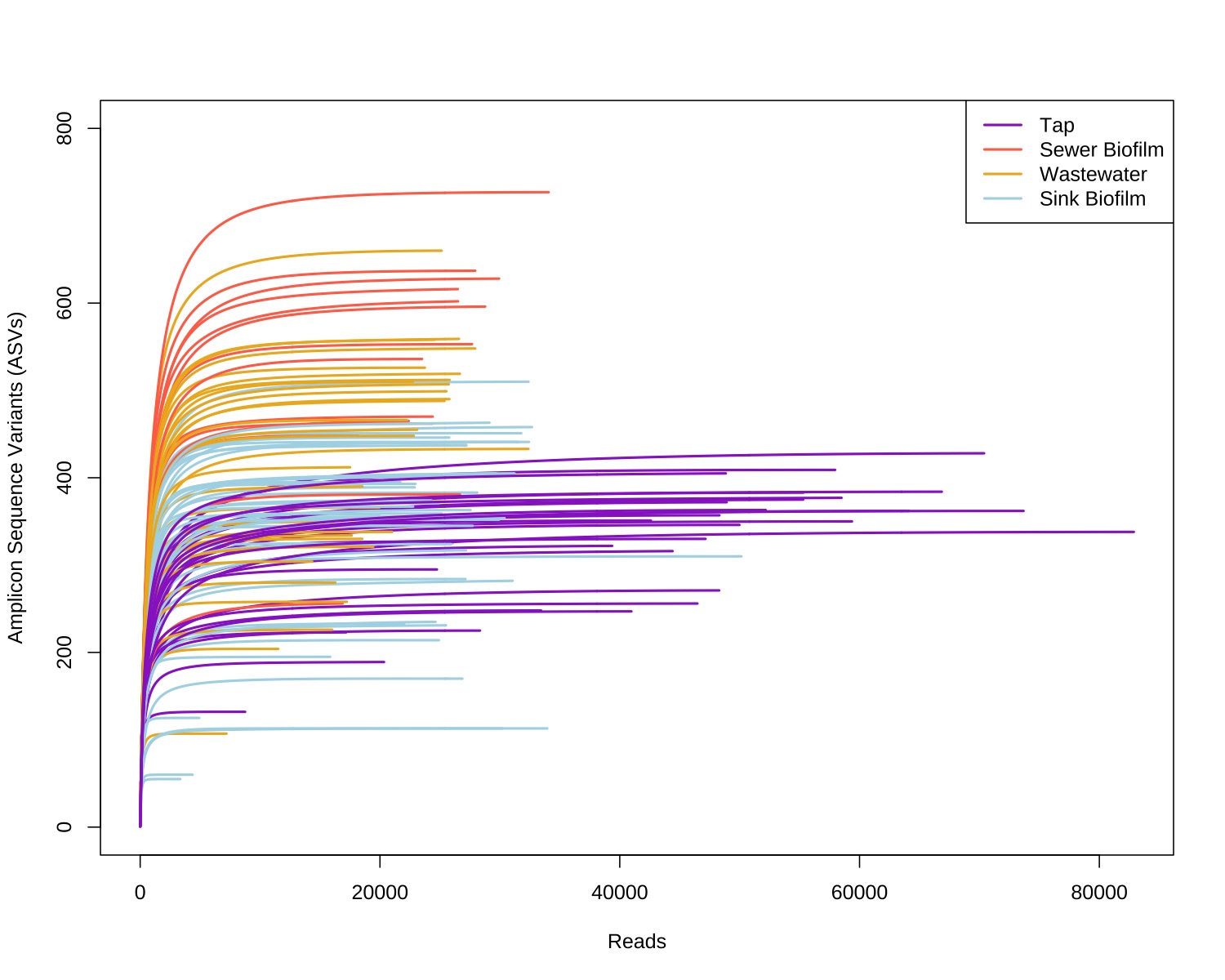

**Figure S2**. Rarefaction curves for the 16S rRNA gene amplicon data with colors representing sample environments.

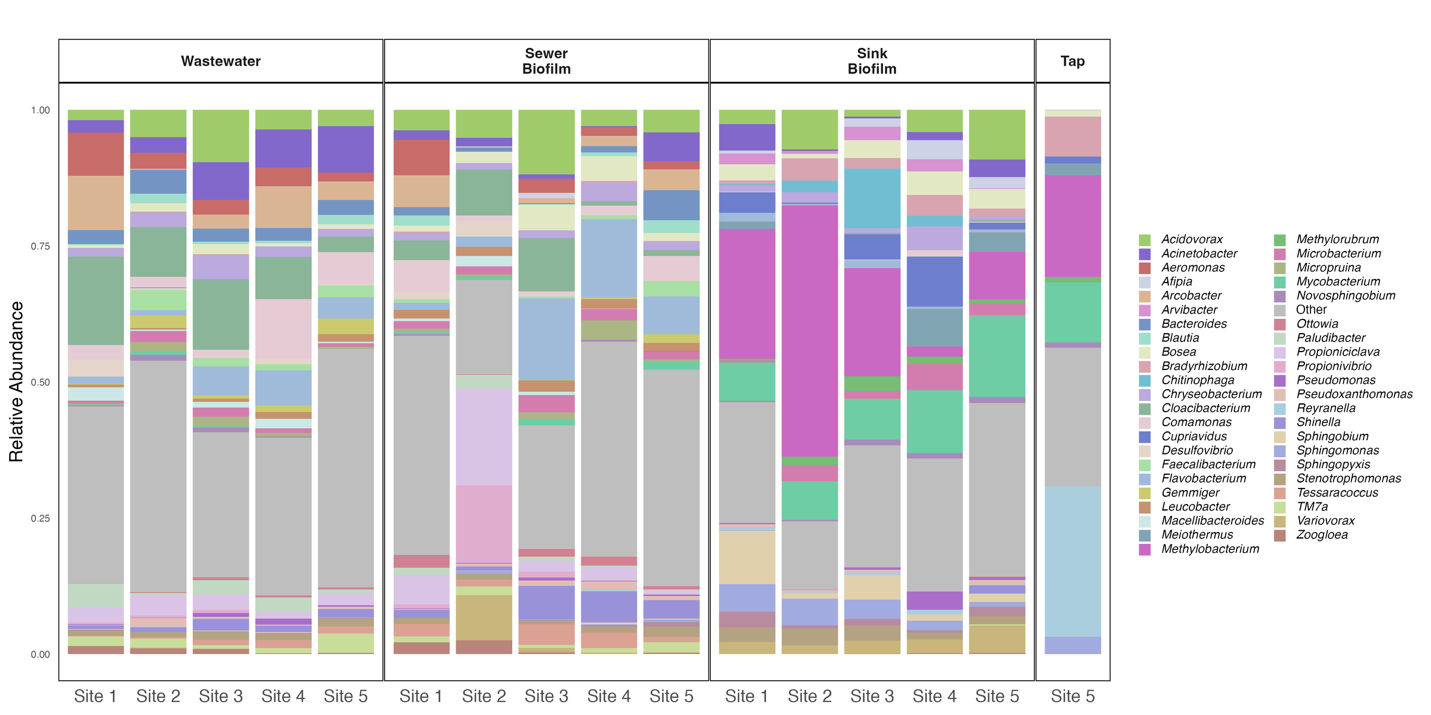

**Figure S3.** **Genus-level community composition across site-associated wastewater, sewer biofilm, sink biofilm, and tap samples, collapsed over time.** Stacked bars show relative abundances of the 44 most abundant genera across Site 1–5 within each sample type; all remaining genera were grouped as “Other.” Counts were agglomerated to genus within each Site × sample-type combination and summed across timepoints (phyloseq merge_samples) before conversion to relative abundance, so each bar represents the pooled community.

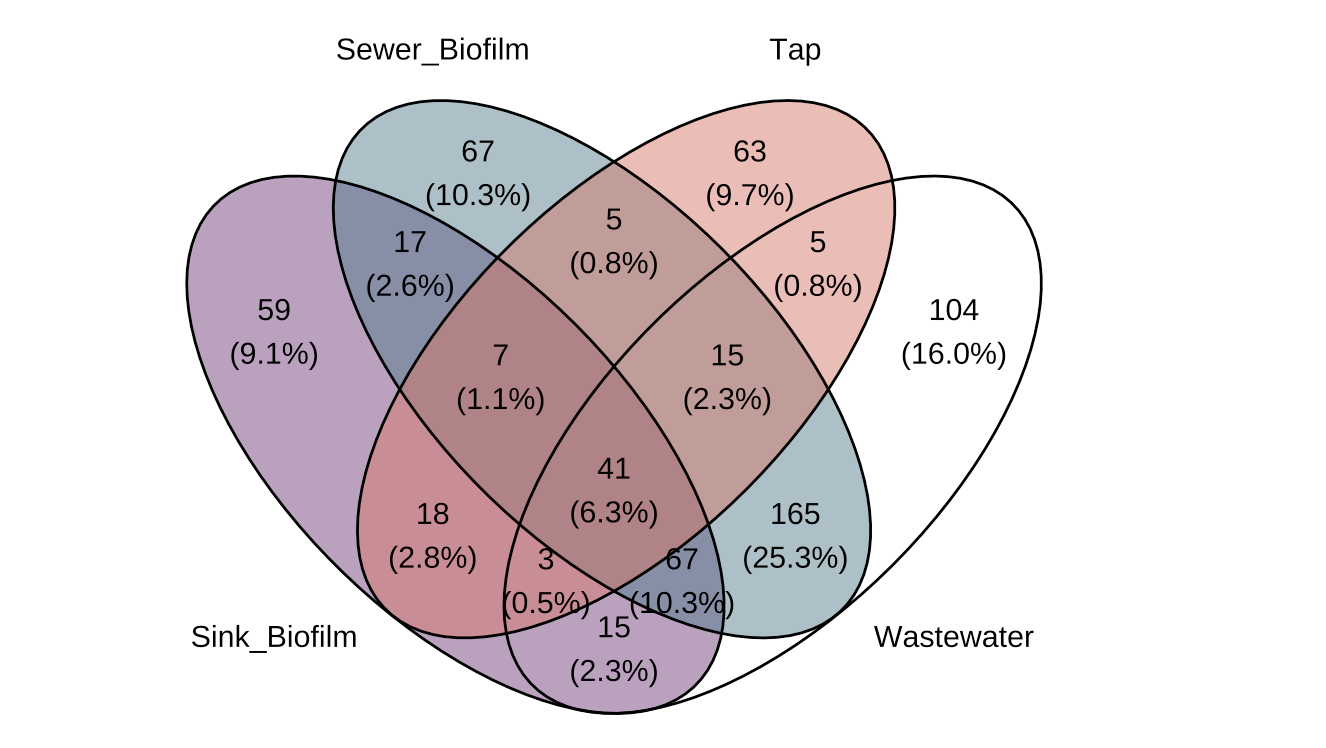

**Figure S4. Overlap in detected bacterial genera across hospital sink drain biofilm, sewer biofilm, and wastewater samples.** Euler diagrams show the shared and unique genera identified by 16S rRNA amplicon sequencing across sample types, based on presence/absence after genus-level taxonomic aggregation. Counts (or percentages) indicate the number (or proportion) of genera detected exclusively within or shared among each environment.

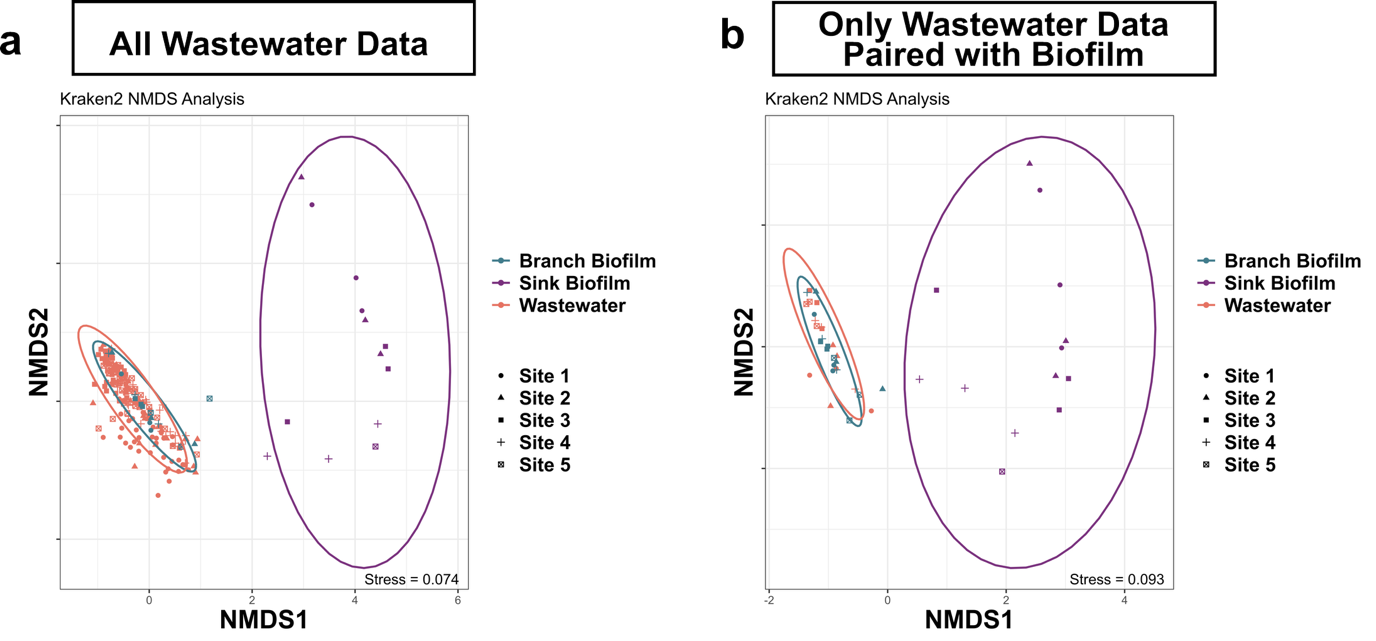

**Figure S5.** **Non-metric multidimensional scaling (NMDS) of probe-capture metagenomics-based pathogen composition across wastewater and biofilm samples.** a) NMDS ordination was performed on Kraken2-derived species-level pathogen profiles for samples with at least one detected pathogen. Community distances were calculated using the Bray-Curtis index. b) **NMDS ordination of RPIP-targeted pathogen communities across hospital sink biofilms, branch biofilms, and temporally matched wastewater samples from five Sites.** Ordination is based on Bray-Curtis dissimilarities of species-level relative abundances from Kraken2 classifications (confidence threshold = 0.15), restricted to RPIP panel taxa. Wastewater samples were matched to the nearest biofilm collection date per Site and timepoint. Ellipses denote 95% confidence regions around each sample-type centroid.

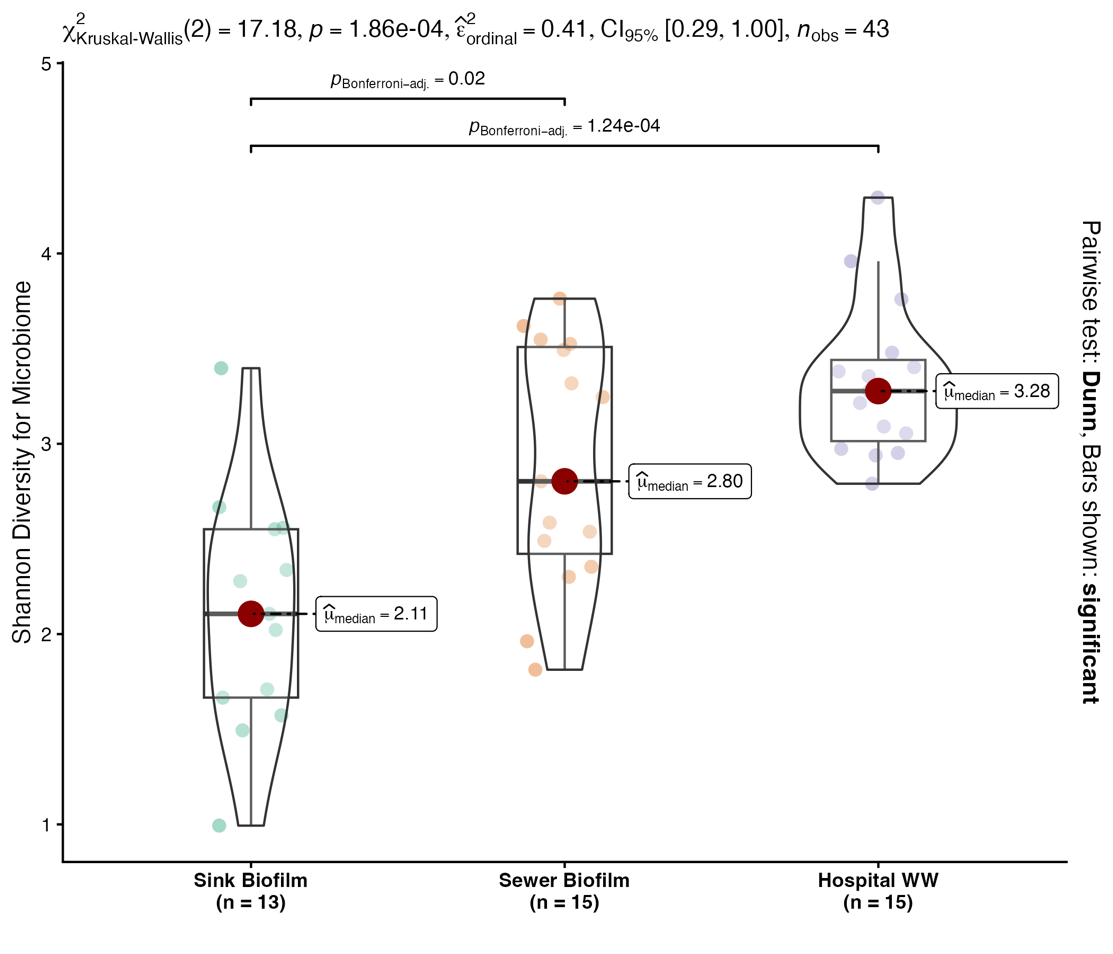

**Figure S6.** Probe-based metagenomic-derived alpha diversity of pathogens across sample types. Shannon diversity was calculated from Kraken2-derived pathogen profiles for hospital wastewater and biofilm samples. Pathogen abundances were presence/absence–binarized and rarefied to standardize sampling depth. Points represent individual samples, bars indicate median Shannon diversity, and sample types on the x-axis are ordered by median diversity. Nonparametric tests with Bonferroni correction were used to assess differences between sample types.

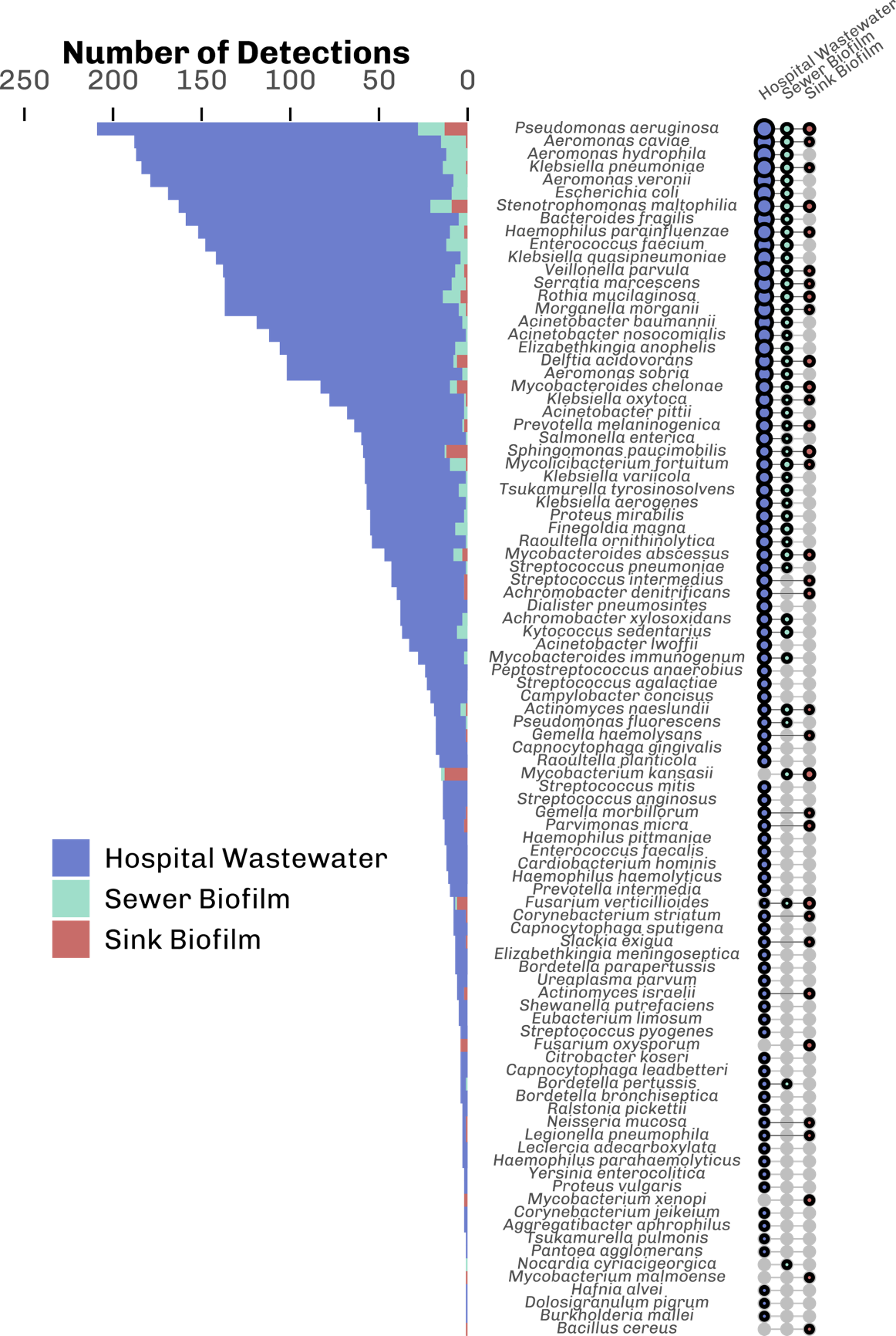

**Figure S7. Detection frequency of pathogens both included and not included in the probe-based metagenomics panel across wastewater and biofilm sample types.** Bar plots show the number of samples in which each pathogen was detected (relative abundance > 0.0005%) across hospital Sites, stratified by sample type (wastewater, sewer biofilm, and sink biofilm). Species are ordered by total detection frequency across all sample types. The accompanying dot plot indicates the distribution of detections across sample types for each species, where dot size corresponds to the number of detections within a given sample type and vertical position denotes sample type. Only pathogens detected at least twice across the study locations are included.

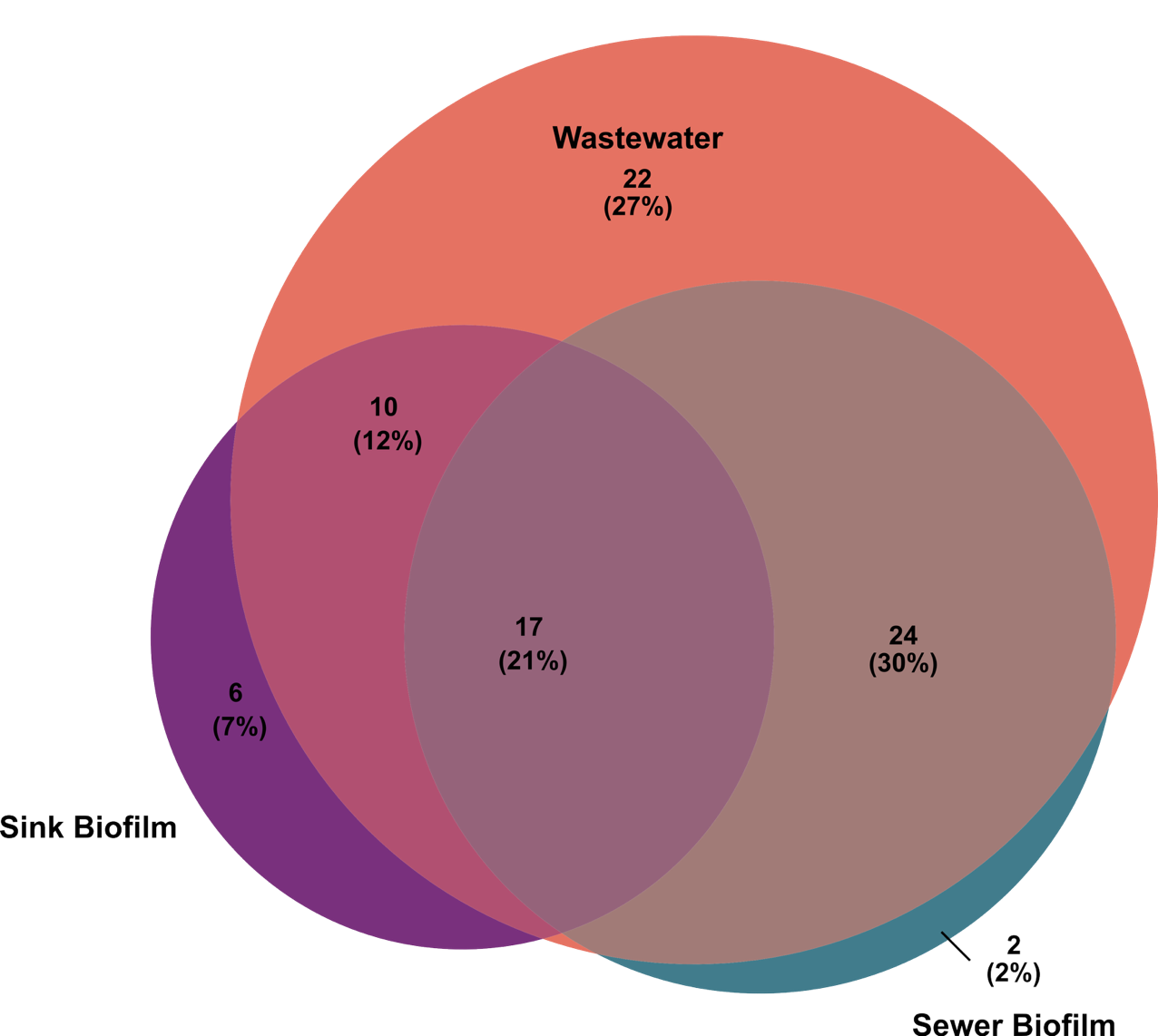

**Figure S8. a) Euler diagram of species-level overlap among sink biofilm, sewer biofilm, and bulk wastewater communities.** Pathogens presence–absence was derived from Kraken2 (confidence-threshold set to 0.5) species-level profiles by retaining taxa with nonzero relative abundance within each sample. For each sampling location (hospital Site), species overlap among sink drain biofilm, sewer biofilm, and bulk wastewater was quantified, and counts for each overlap region were calculated. Overlap counts were then averaged across locations to generate a composite representation of shared and unique taxa. The Euler diagram displays the mean number (or percentage) of species unique to each sample type and shared between sample types, illustrating the extent of community overlap between biofilm-associated and wastewater microbiomes.

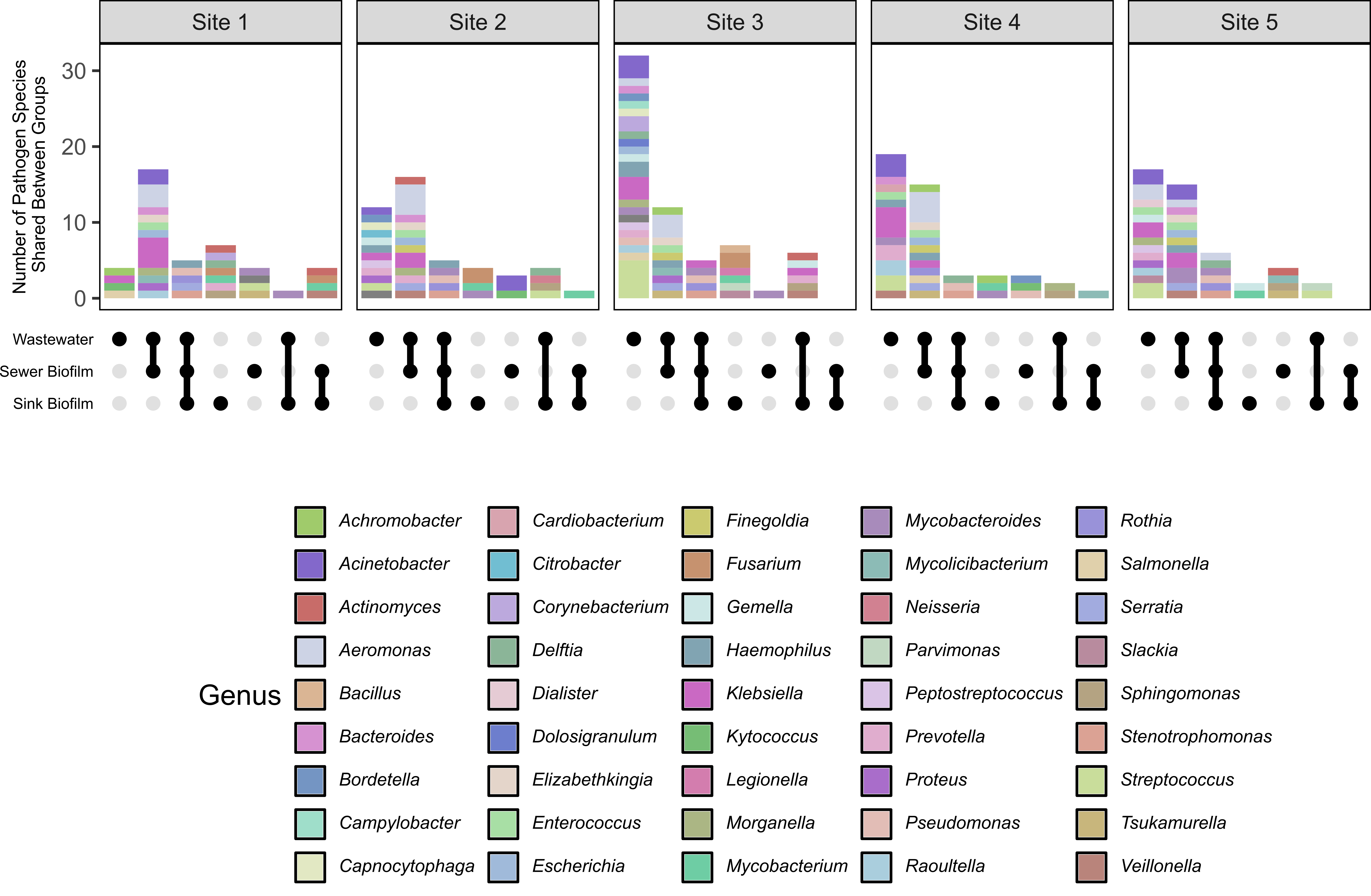

**Figure S9. Genus-level overlap of RPIP pathogen detections across sink biofilm, sewer biofilm, and hospital wastewater, faceted by Site.** UpSet plots depict the presence–absence overlap of RPIP pathogen genera detected at least once within each environment and Site. Genus-level detections were derived from species-level Kraken2 annotations, with presence defined as detection in any sample from a given environment. Bars represent the number of pathogens shared across combinations of sample types, colored by genus (top-ranked genera shown; remaining grouped as “Other”). Site-level faceting highlights differences in genus sharing across biofilm-associated and wastewater environments.

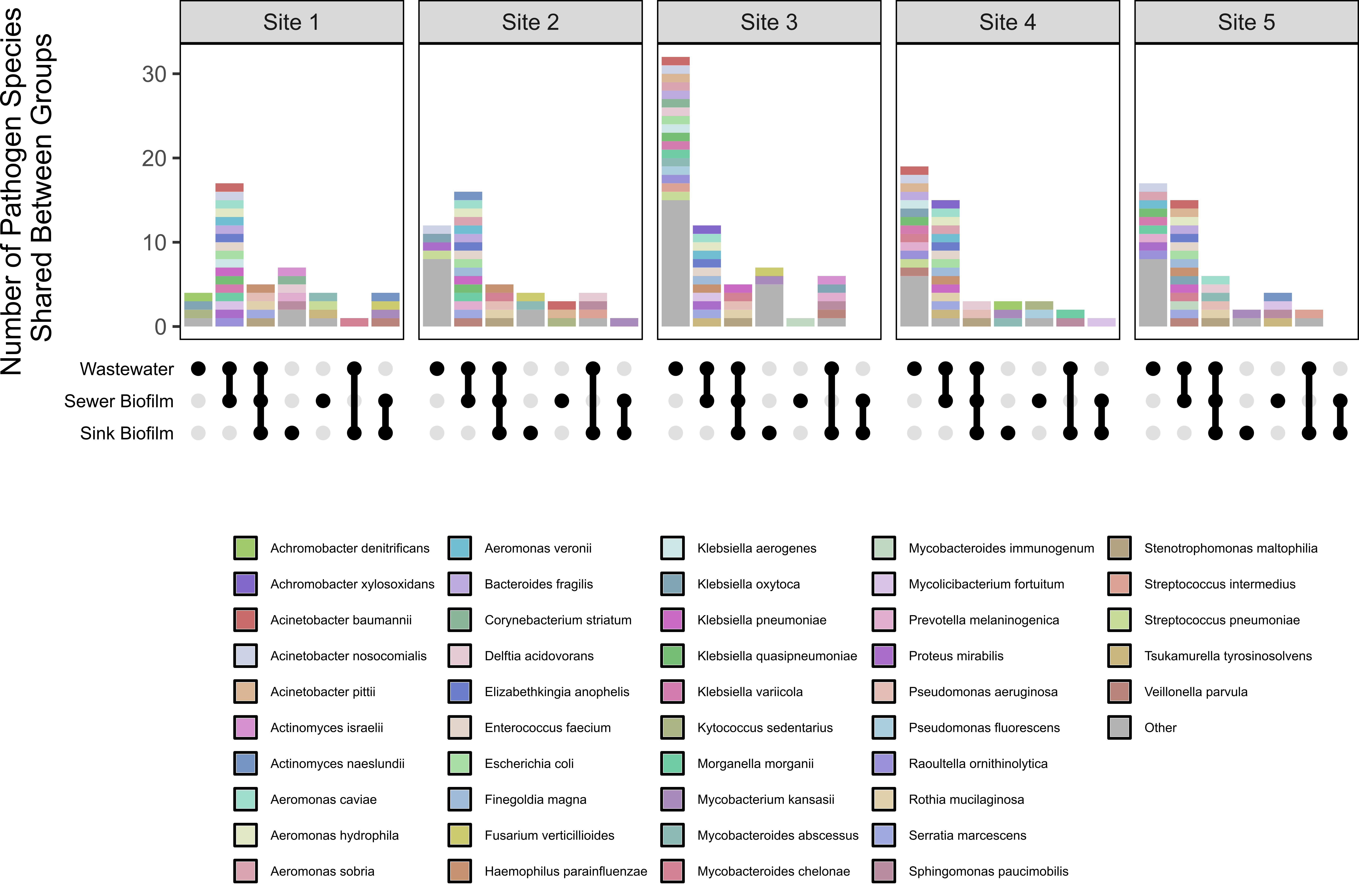

**Figure S10. Species-level overlap of RPIP pathogen detections across sink biofilm, sewer biofilm, and hospital wastewater, faceted by Site.** UpSet plots summarize the presence–absence overlap of RPIP pathogen species detected at least once in each environment within a Site. Species detections were derived from Kraken2 profiles and binarized such that a species was considered present if detected in any sample from a given environment. Bars indicate the number of pathogen species observed in each combination of sample types, with colors denoting individual species (top-ranked species shown; remaining species grouped as “Other”). Facets correspond to hospital Sites.

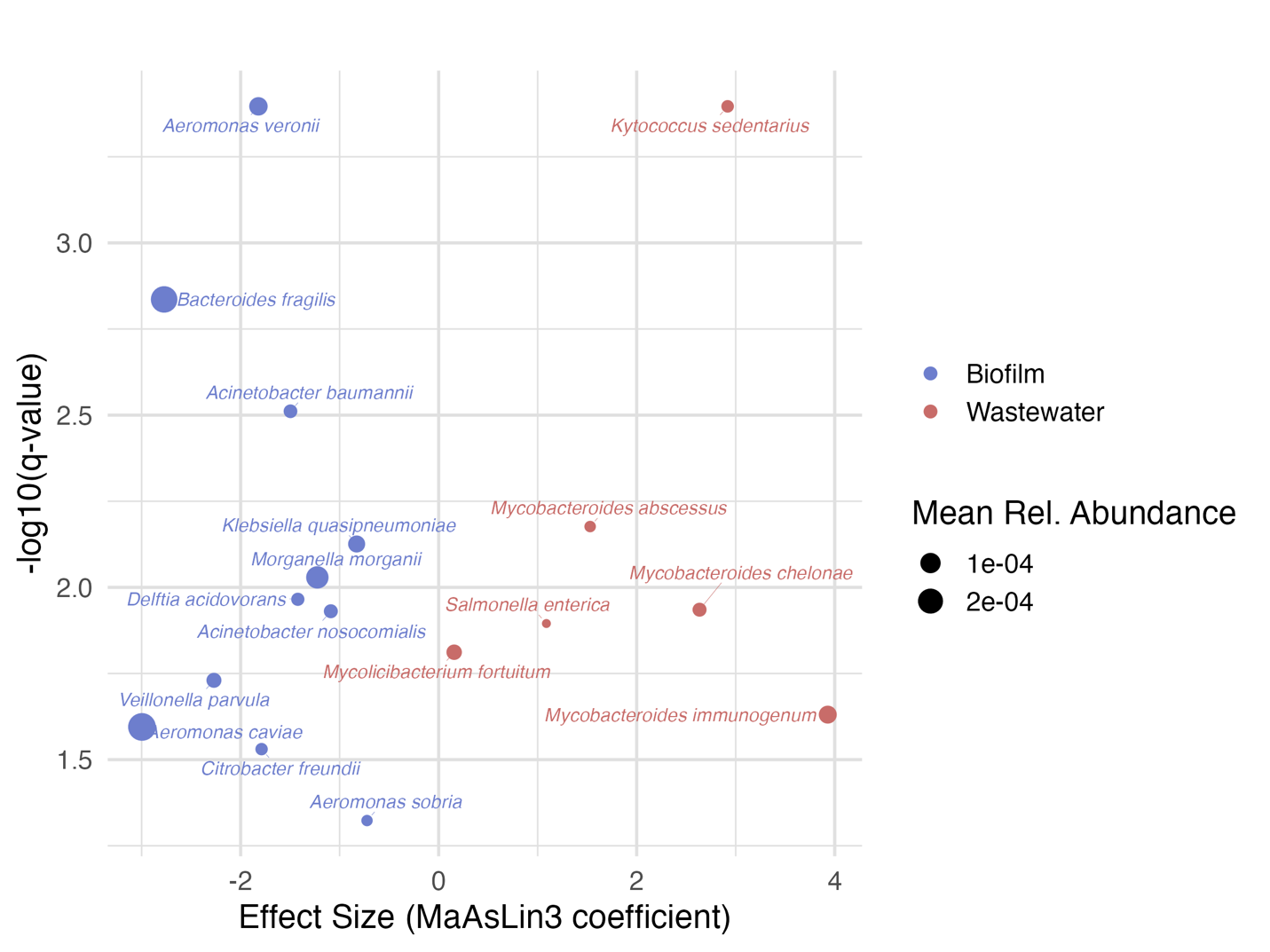

**Figure S11. Volcano plot of differentially abundant RPIP pathogens between sewer biofilm and wastewater samples identified using MaAsLin3.** Each point represents a pathogen detected at species level, with the MaAsLin3 coefficient (effect size) on the x-axis and statistical significance shown as −log10(q-value) on the y-axis. Positive coefficients indicate enrichment in wastewater, while negative coefficients indicate enrichment in biofilm. Point size reflects mean relative abundance across samples, and labeled taxa denote the most significant or abundant pathogens. Analyses were restricted to temporally matched sampling dates shared between biofilm and wastewater across locations, with hospital Site included as a random effect.

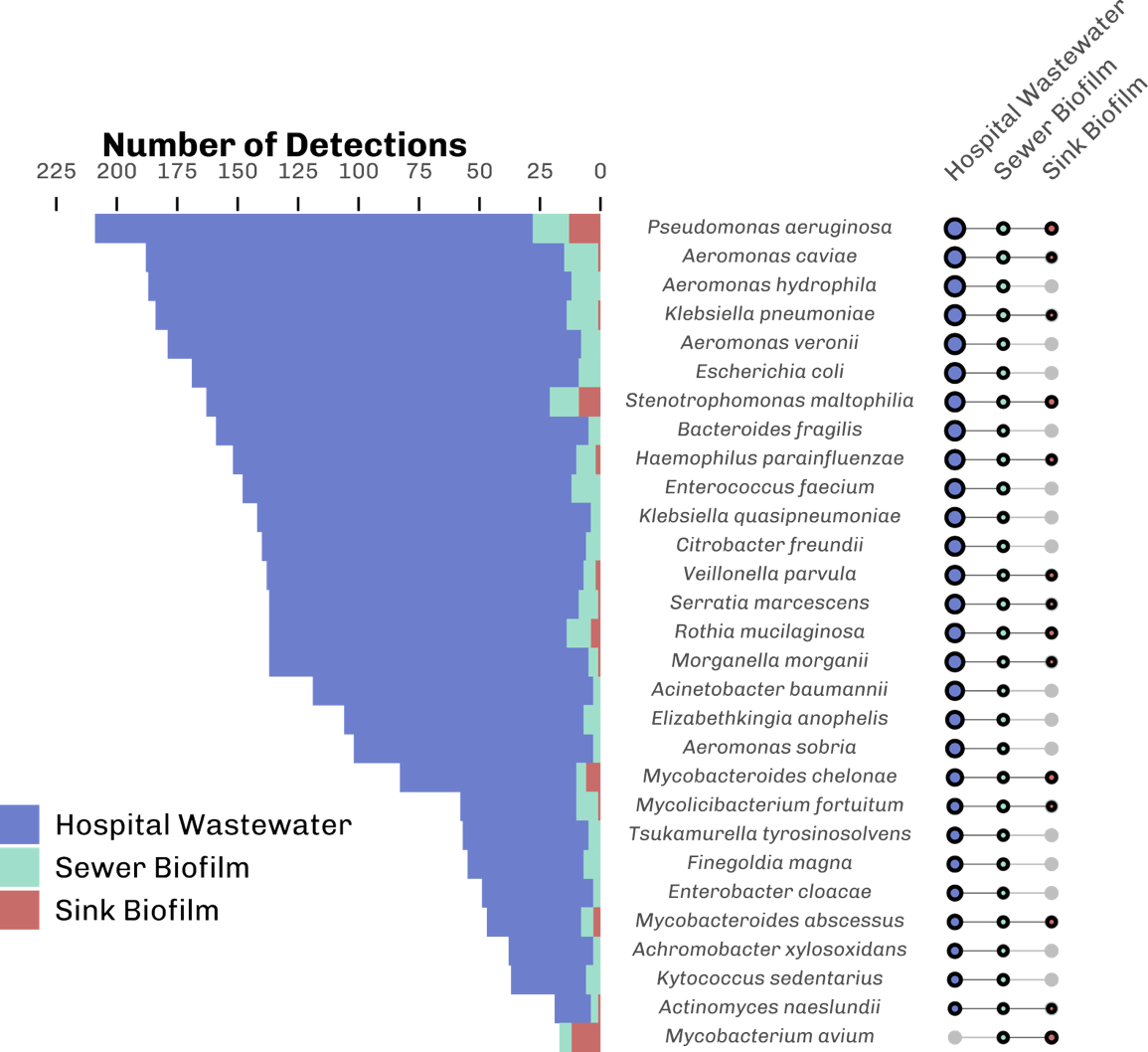

**Figure S12. Pathogen detection patterns across hospital wastewater, sewer biofilm, and sink biofilm.** Shown are RPIP target pathogen species identified by Kraken2 that were detected in at least three sewer biofilm samples. Horizontal bars represent the total number of detections per species across all sample types, and the adjacent dot plot shows the distribution of detections within hospital wastewater, sewer biofilm, and sink biofilm. Larger points indicate greater detection frequency within that environment. Species are ranked by total detections across all environments.

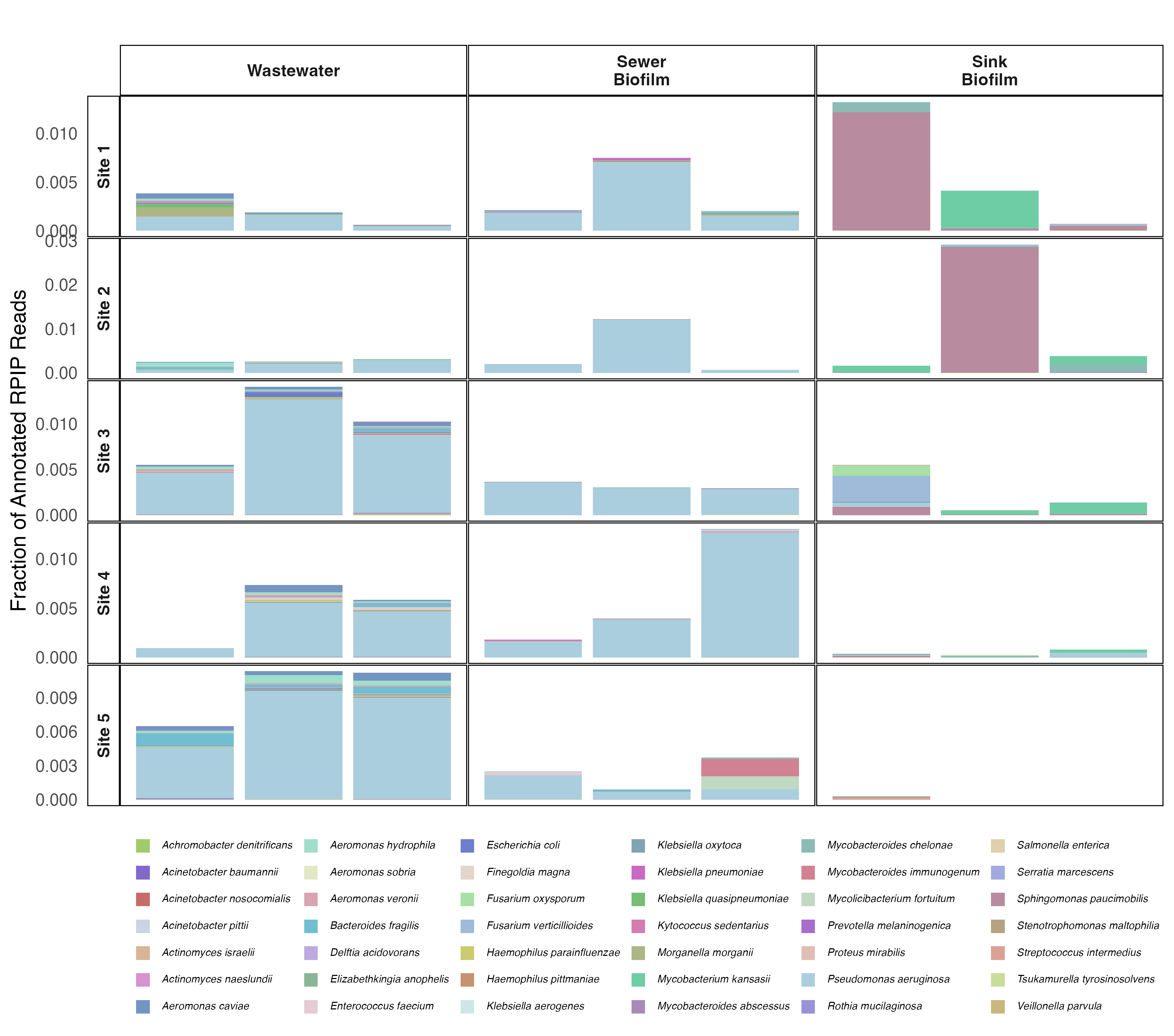
**Figure S13.** Relative abundance of RPIP-target taxa across paired hospital wastewater and biofilm samples collected from five Sites between September–December 2024. Stacked bars show the fraction of reads assigned to RPIP pathogen species (species-level Kraken annotations) for each sample, faceted by site (rows) and sample type (columns: wastewater, sewer biofilm, sink biofilm). To harmonize sampling across matrices, wastewater and sink biofilm samples were temporally matched to the corresponding sewer biofilm collection date within each Site/timepoint. The 42 most abundant RPIP species (by summed relative abundance) across all samples are shown individually; remaining taxa are grouped as or “Other Enriched Pathogens.” Non-enriched taxa were removed from the plot.

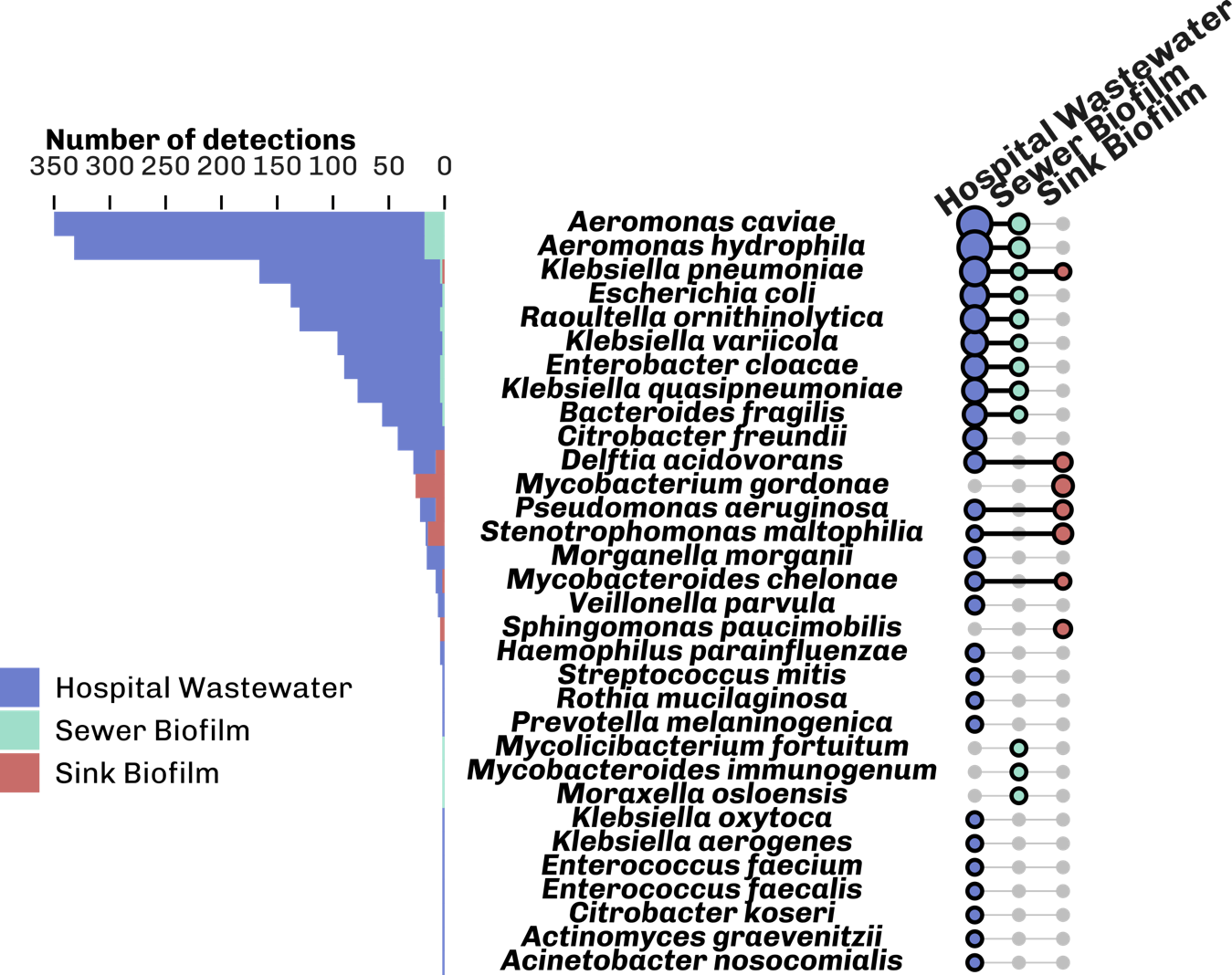

**Figure S14. Detection frequency of RPIP pathogen species detected using MetaPhlAn across hospital wastewater and biofilm sample types.** Bar plots show the number of samples in which each RPIP pathogen species was detected (relative abundance > 0) across hospital Sites, stratified by sample type (hospital wastewater, sewer biofilm, and sink biofilm). Species are ordered by total detection frequency across all sample types. The accompanying dot plot indicates the distribution of detections across sample types for each species, where dot size corresponds to the number of detections within a given sample type and vertical position denotes sample type. Only RPIP target species detected at least once across the study locations are included.

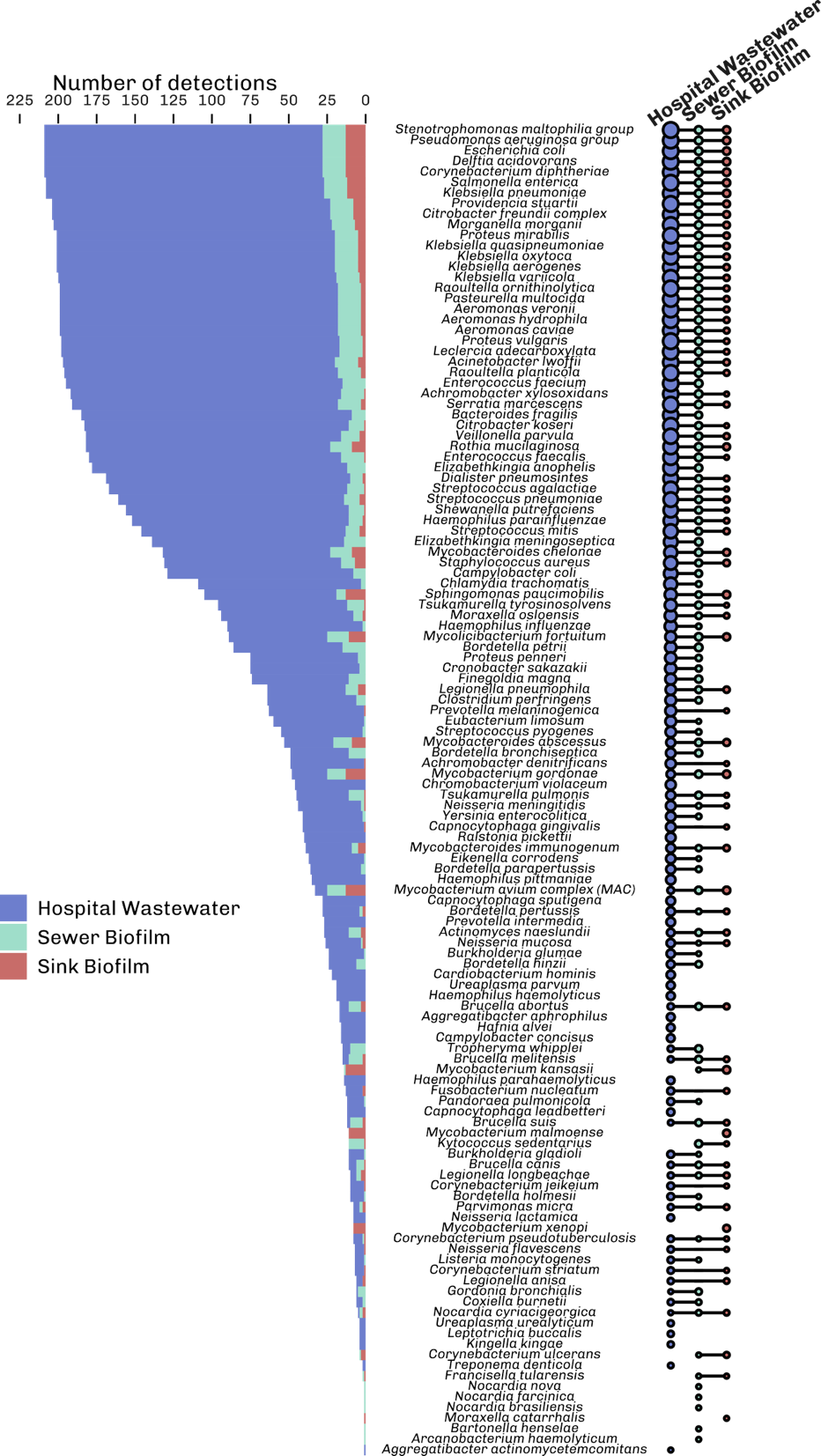

**Figure S15. Detection frequency of RPIP pathogen species detected using Centrifuger across hospital wastewater and biofilm sample types.** Bar plots show the number of samples in which each RPIP pathogen species was detected (relative abundance > 0) across hospital Sites, stratified by sample type (hospital wastewater, sewer biofilm, and sink biofilm). Species are ordered by total detection frequency across all sample types. The accompanying dot plot indicates the distribution of detections across sample types for each species, where dot size corresponds to the number of detections within a given sample type and vertical position denotes sample type. Only RPIP target species detected at least once across the study locations are included.

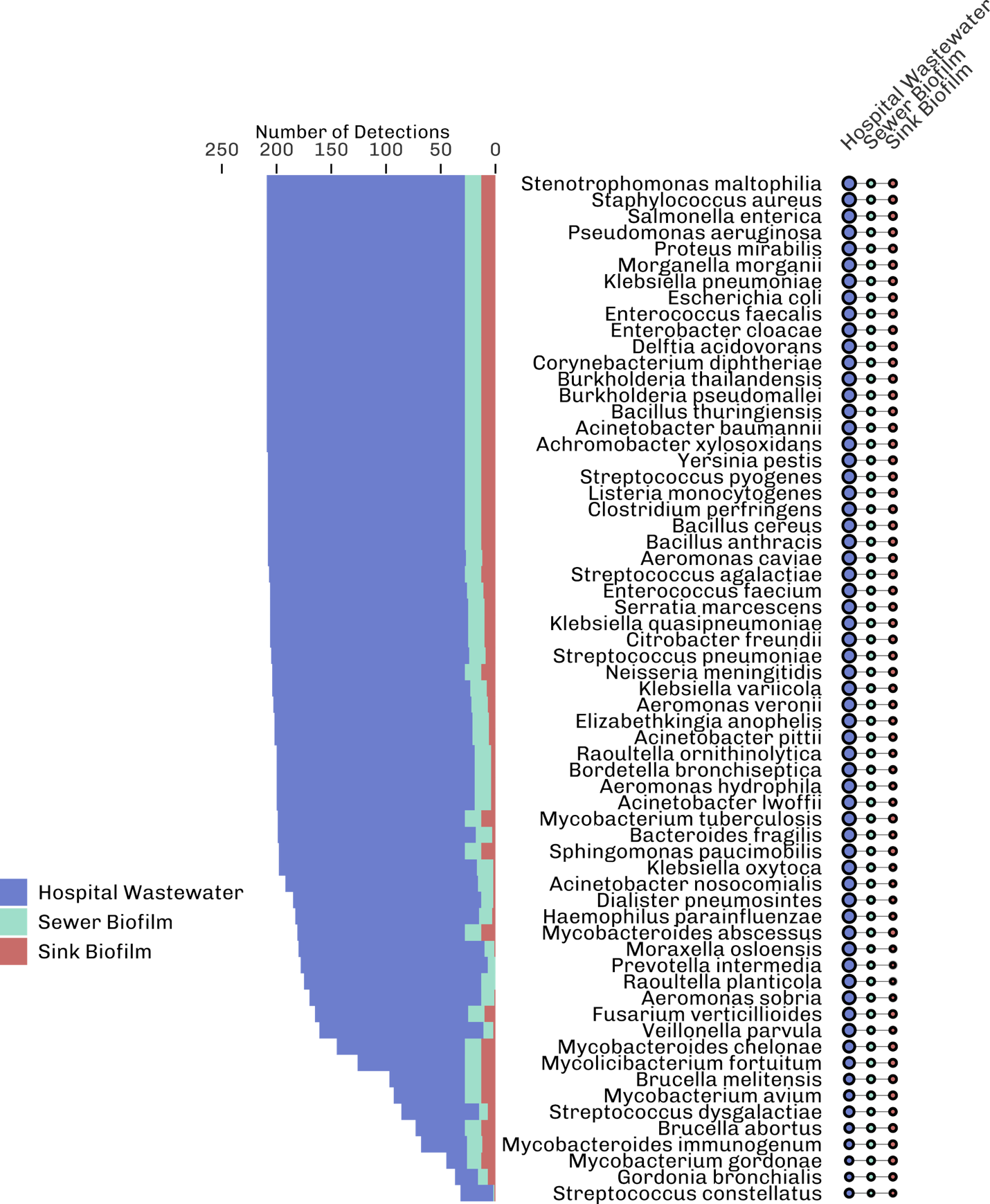

**Figure S16. Detection frequency of RPIP pathogen species detected using Kraken2 (CT: 0) across hospital wastewater and biofilm sample types.** Bar plots show the number of samples in which each RPIP pathogen species was detected (relative abundance > 0) across hospital Sites, stratified by sample type (hospital wastewater, sewer biofilm, and sink biofilm). Species are ordered by total detection frequency across all sample types. The accompanying dot plot indicates the distribution of detections across sample types for each species, where dot size corresponds to the number of detections within a given sample type and vertical position denotes sample type. Only RPIP target species detected at least once across the study locations are included.

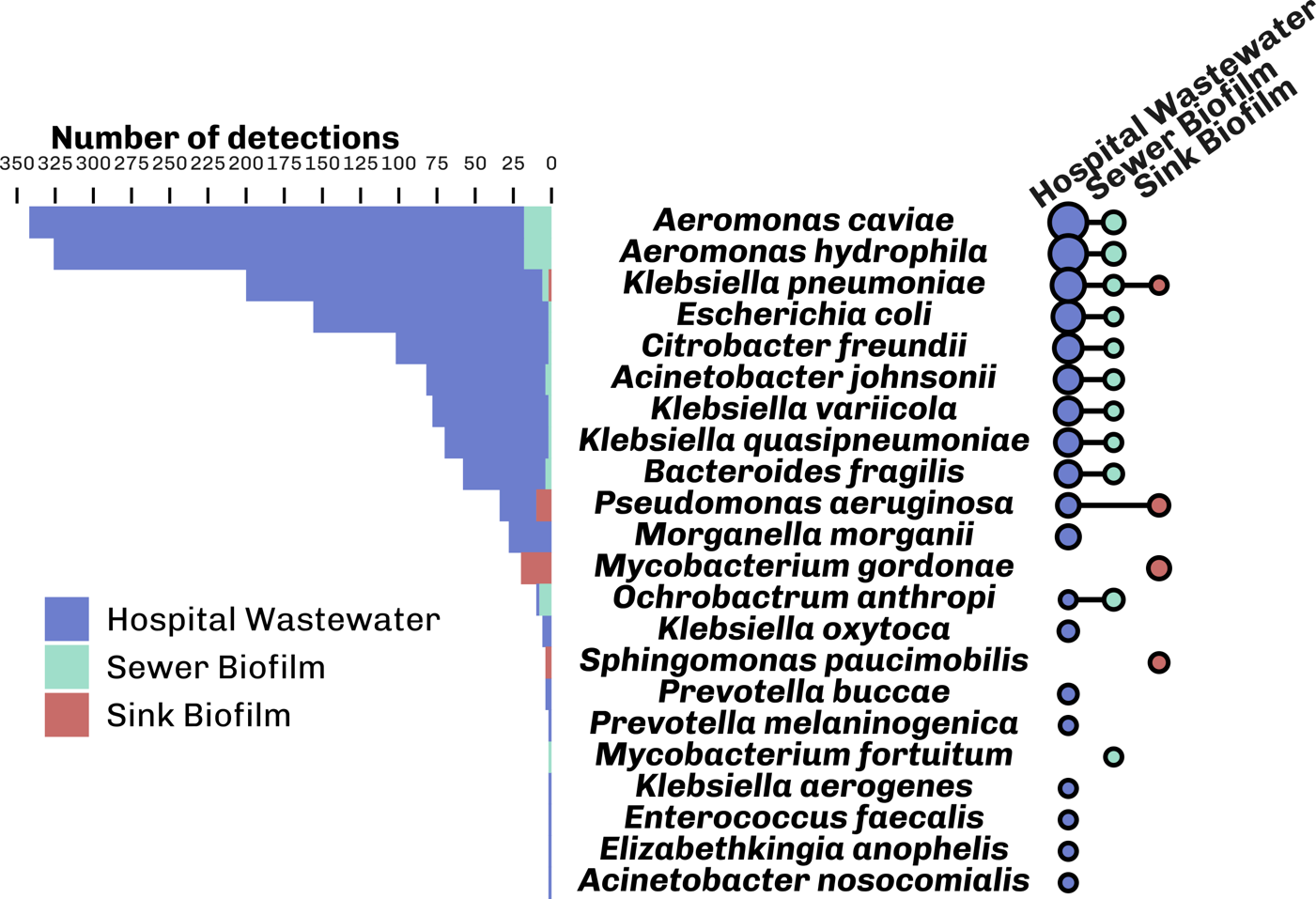

**Figure S17. Detection frequency of RPIP pathogen species detected using Sylph across hospital wastewater and biofilm sample types.** Bar plots show the number of samples in which each RPIP pathogen species was detected (relative abundance > 0) across hospital Sites, stratified by sample type (hospital wastewater, sewer biofilm, and sink biofilm). Species are ordered by total detection frequency across all sample types. The accompanying dot plot indicates the distribution of detections across sample types for each species, where dot size corresponds to the number of detections within a given sample type and vertical position denotes sample type. Only RPIP target species detected at least once across the study locations are included.

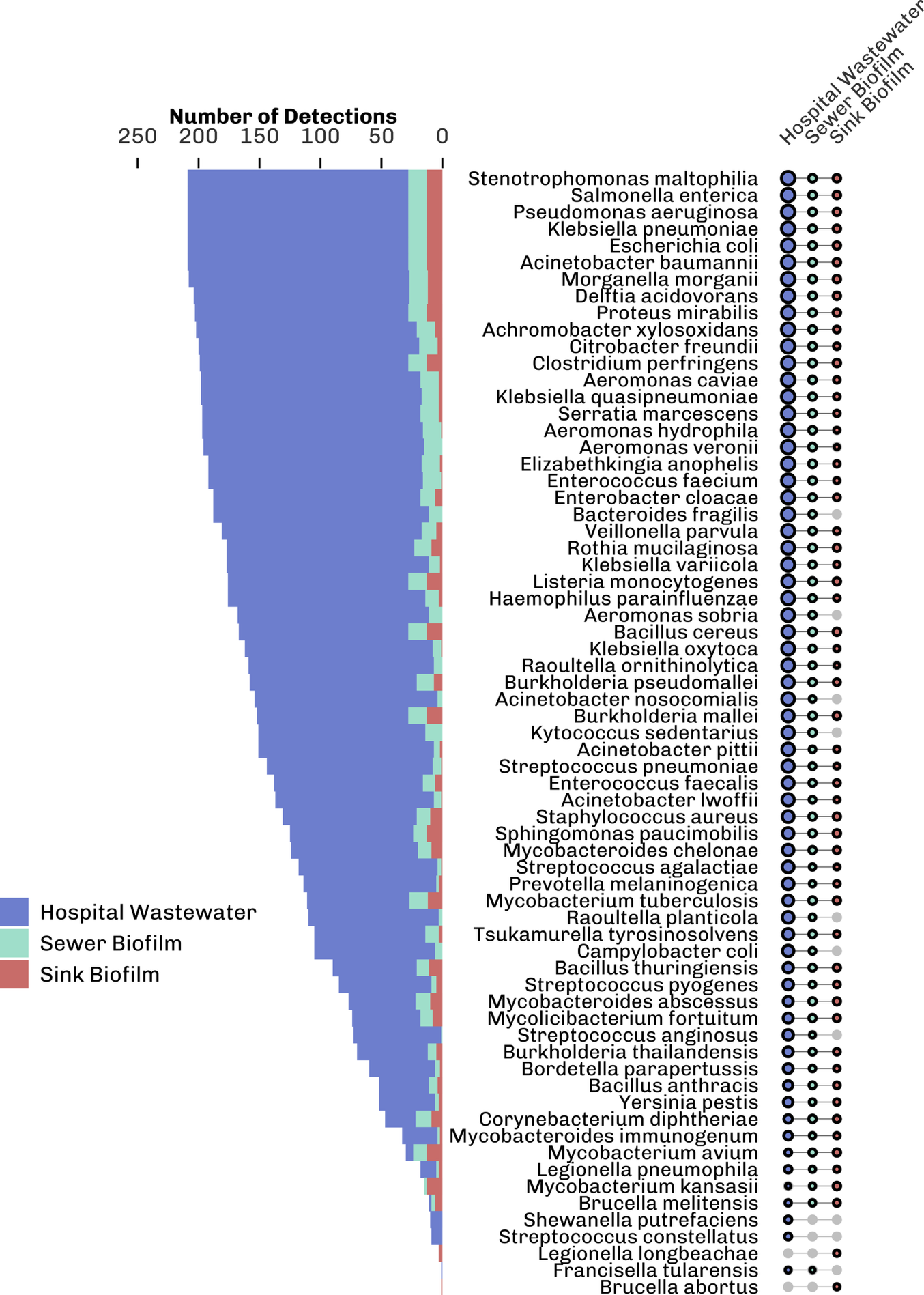

**Figure S18. Detection frequency of RPIP pathogen species detected using Kraken2 (CT: 0.2) across hospital wastewater and biofilm sample types.** Bar plots show the number of samples in which each RPIP pathogen species was detected (relative abundance > 0) across hospital Sites, stratified by sample type (hospital wastewater, sewer biofilm, and sink biofilm). Species are ordered by total detection frequency across all sample types. The accompanying dot plot indicates the distribution of detections across sample types for each species, where dot size corresponds to the number of detections within a given sample type and vertical position denotes sample type. Only RPIP target species detected at least once across the study locations are included.

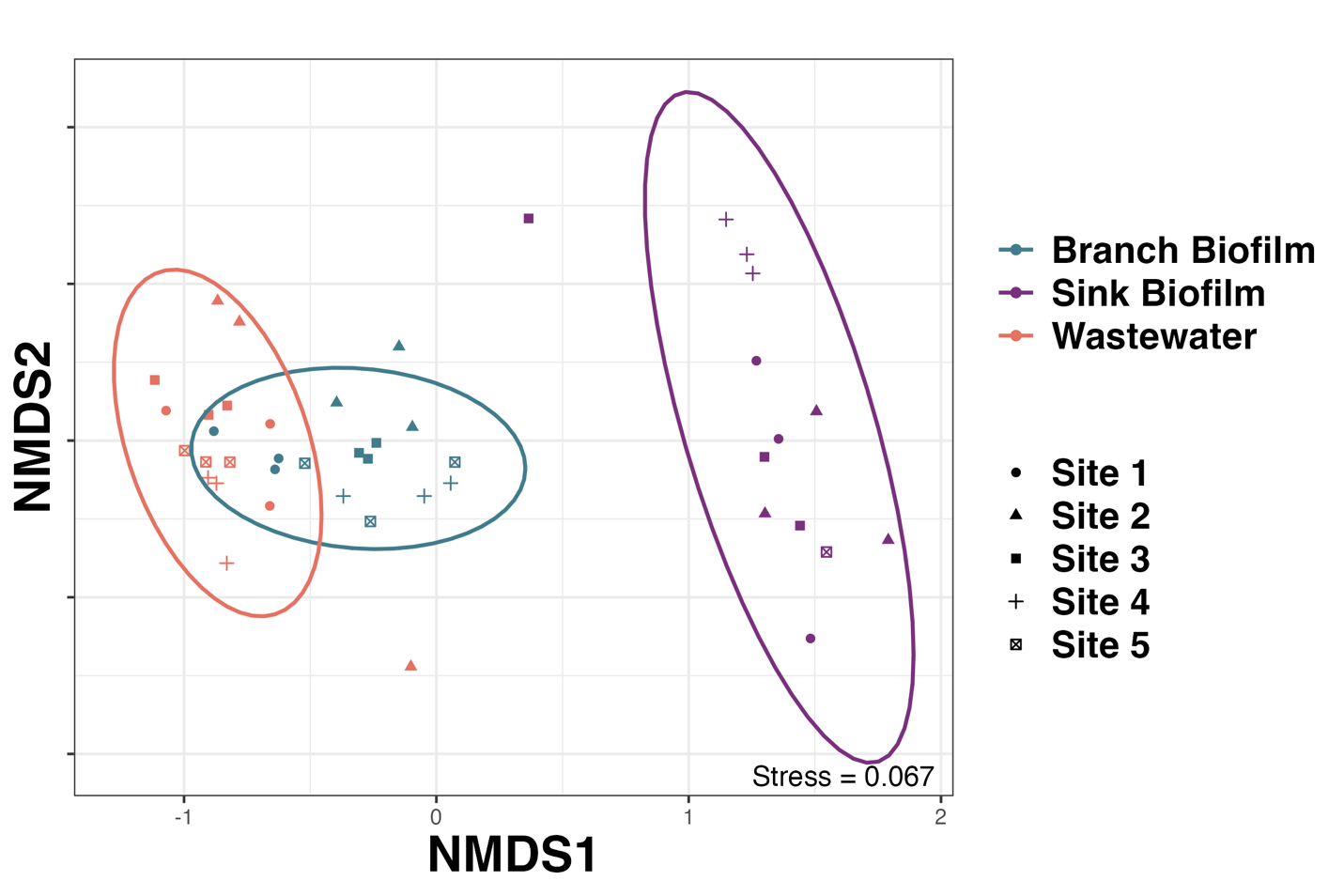

**Figure S19.** **NMDS of resistome community composition across wastewater and biofilm samples.** NMDS ordination was performed on CARD-derived ARG profiles for samples with at least one detected ARG. ARG relative abundances were aggregated per sample and used to compute between-sample dissimilarity. Community distances were calculated using the Bray-Curtis index, emphasizing differences in species presence–absence rather than relative abundance. Each point represents an individual sample, colored by sample type and shaped by location, with ellipses indicating the dispersion of samples within each group.

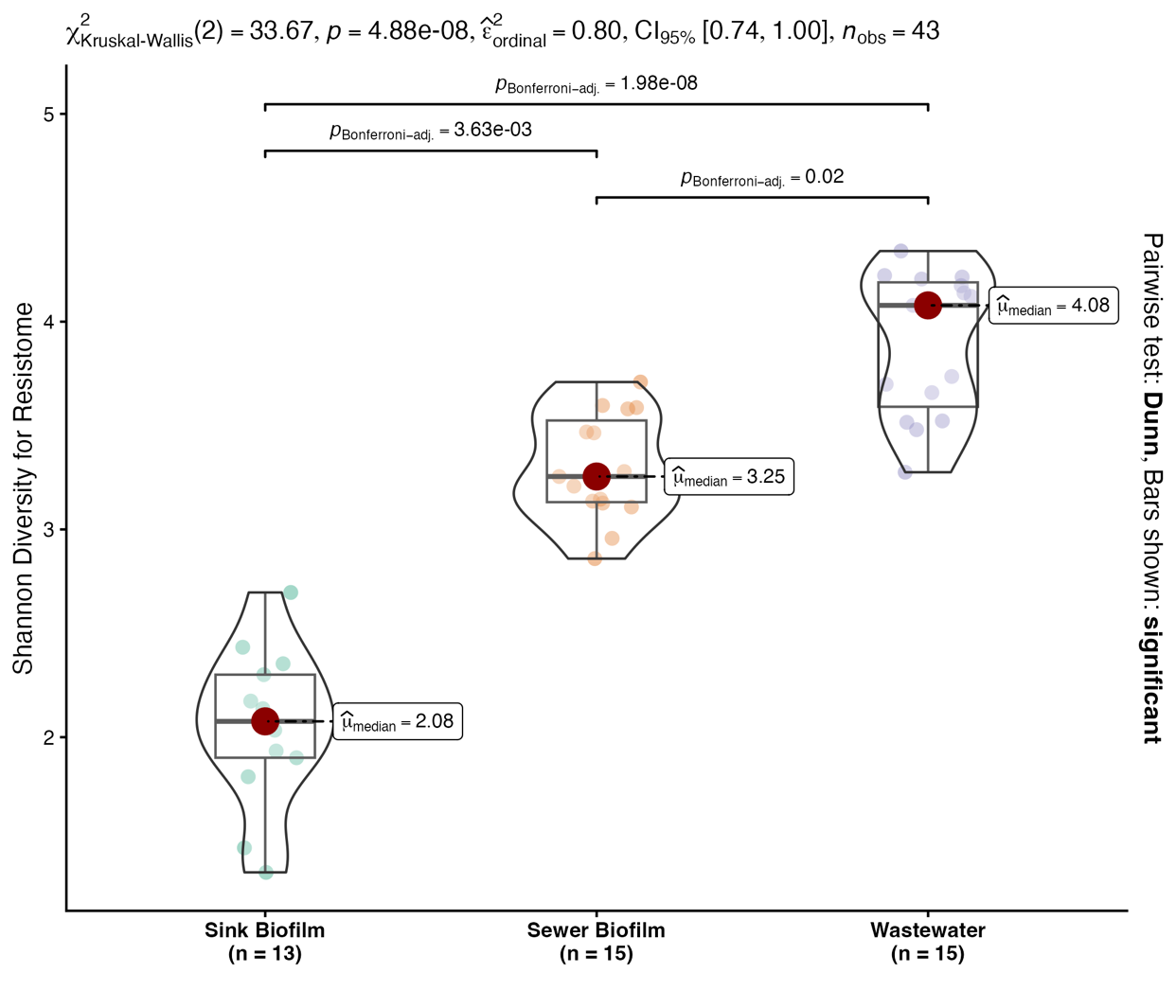

**Figure S20.** Resistome alpha diversity across sample types. Shannon diversity was calculated from CARD-derived ARG profiles for hospital wastewater and biofilm samples. ARG abundances were presence/absence–binarized and rarefied to standardize sampling depth. Points represent individual samples, bars indicate median Shannon diversity, and sample types on the x-axis are ordered by median diversity. Nonparametric tests with Bonferroni correction were used to assess differences between sample types.

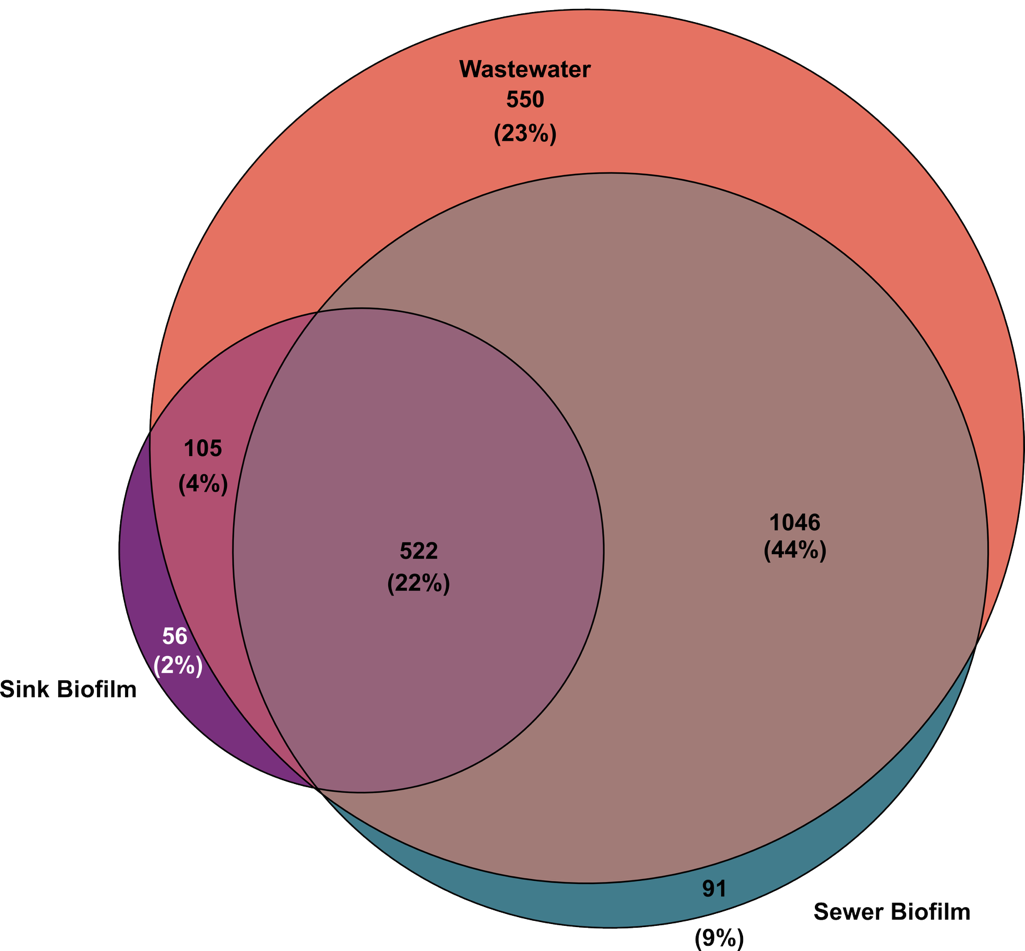

**Figure S21. Euler diagram of ARG overlap among sink biofilm, sewer biofilm, and bulk wastewater communities.** ARG presence–absence was derived from CARD ARG profiles by retaining taxa with nonzero relative abundance within each sample. For each sampling location (hospital Site), species overlap among sink drain biofilm, sewer biofilm, and bulk wastewater was quantified, and counts for each overlap region were calculated. Overlap counts were then averaged across locations to generate a composite representation of shared and unique taxa. The Euler diagram displays the mean number (or percentage) of ARGs unique to each sample type and shared between sample types, illustrating the extent of community overlap between biofilm-associated and wastewater resistomes.

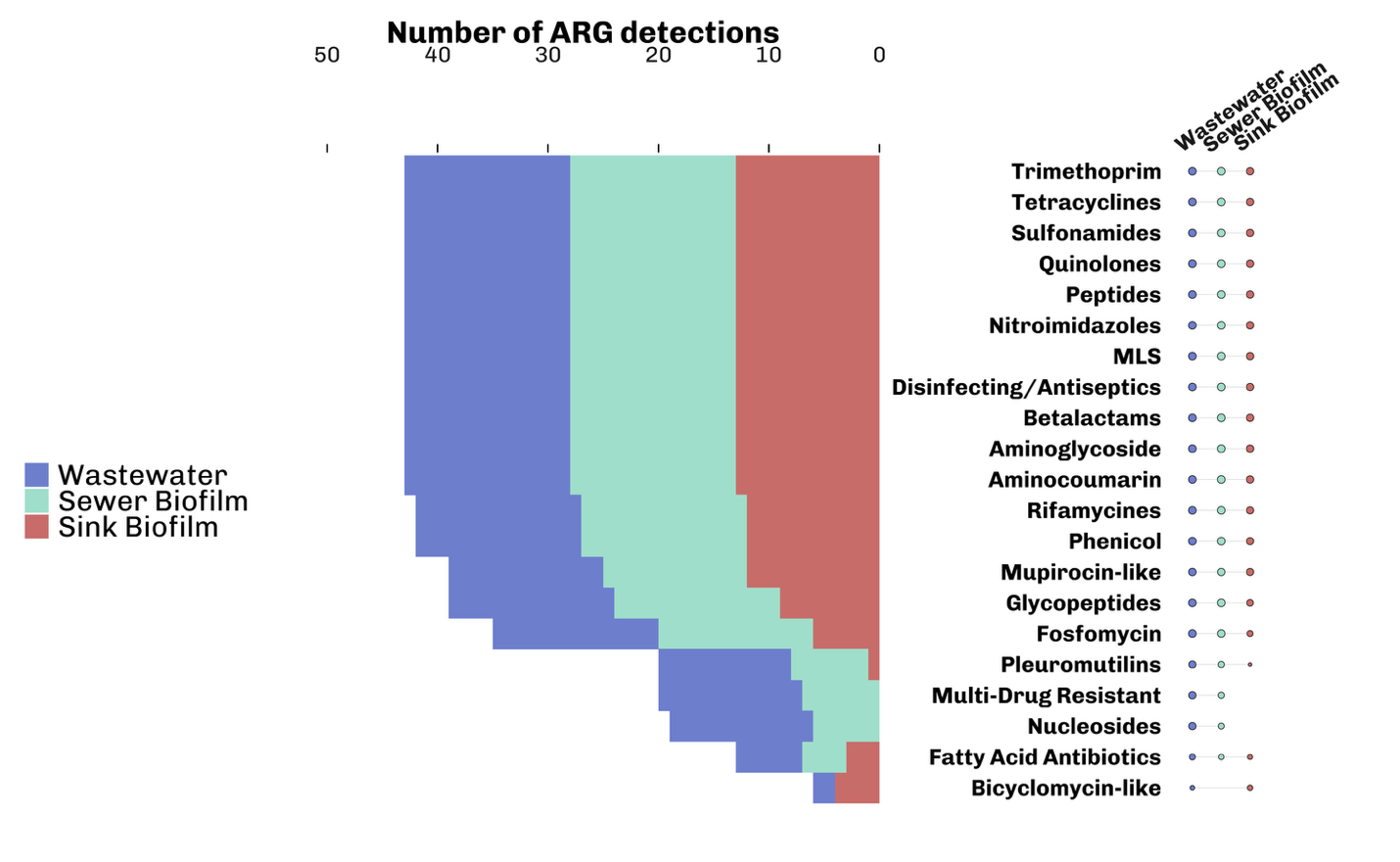

**Figure S22. Detection frequency of AMR Drug Classes detected using DIAMOND and aligned to CARD across hospital wastewater and biofilm sample types.** Bar plots show the number of samples in which each AMR drug class was detected (relative abundance > 0) across hospital Sites, stratified by sample type (hospital wastewater, sewer biofilm, and sink biofilm). Species are ordered by total detection frequency across all sample types. The accompanying dot plot indicates the distribution of detections across sample types for each class, where dot size corresponds to the number of detections within a given sample type and vertical position denotes sample type. Only AMR drug classes detected at least once across the study locations are included.

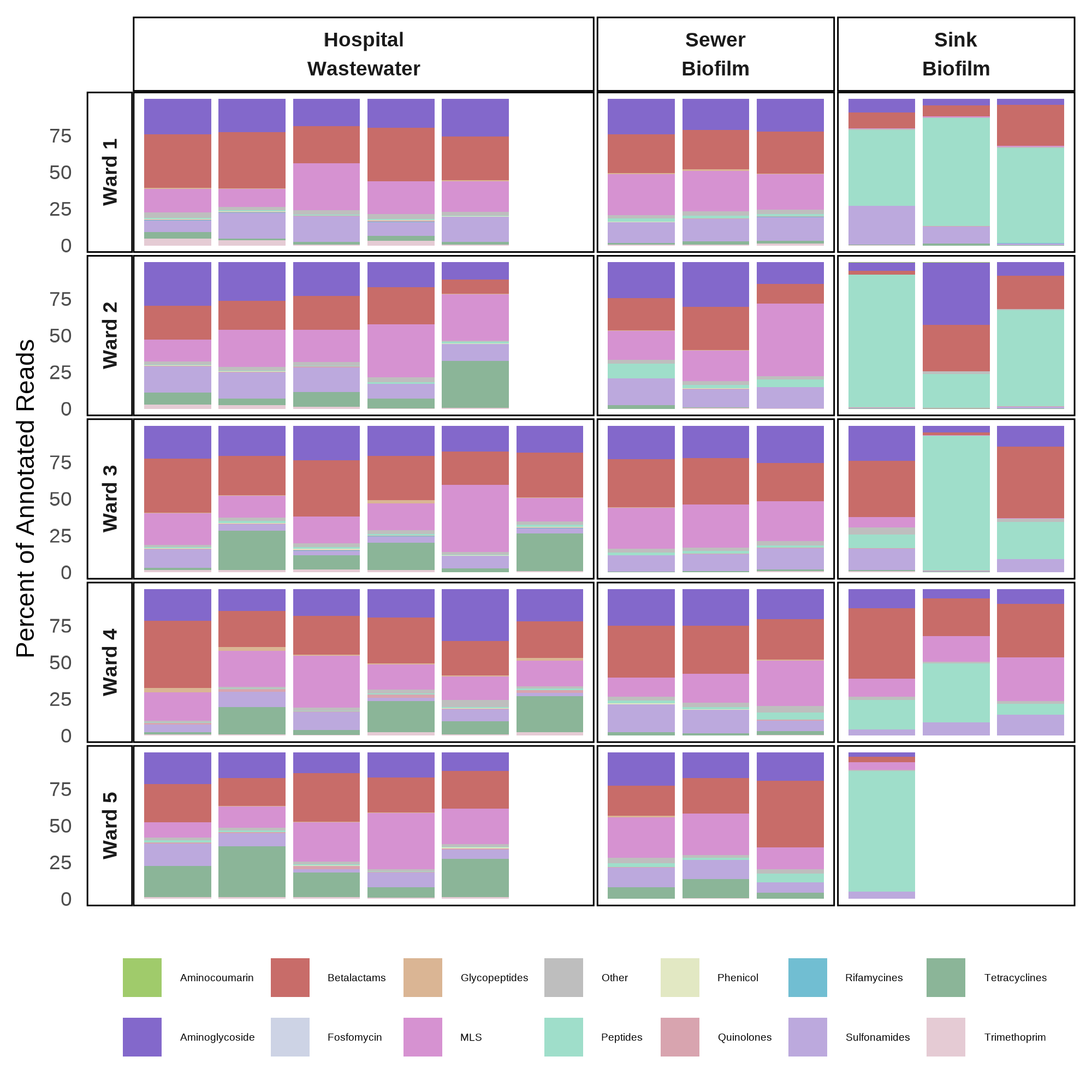

**Figure S23.** Relative composition of ARGs across sink biofilm, sewer biofilm, and hospital wastewater samples collected on shared sampling dates. Stacked bar plots show the relative abundance of ARGs detected by CARD alignment in individual samples, restricted to wastewater collection dates for which biofilm samples were collected immediately prior to or following. Samples are ordered by collection date within each environment and Site, with each bar representing a unique sample–date combination (x-axis index). Facets are nested by site and sample type (sink biofilm, sewer biofilm, hospital wastewater). The most abundant Drug Classes are shown individually, with remaining drug classes grouped as “Other.
